# TorchGWAS2: Cost-Effective Phenome- and Genome-Wide Association Testing in Related Samples

**DOI:** 10.64898/2026.09.10.26362744

**Authors:** Mengyu Zhang, Ziqian Xie, Samaneh Salehi Nasab, Nannan Wang, Xingzhong Zhao, Taryn Alkis, John Barnard, Thomas W. Blackwell, Russell P. Bowler, Shinhye Chung, Michael H. Cho, Clary B. Clish, Emily Drzymalla, Anne M. Evans, Nora Franceschini, Robert E. Gerszten, Madeline G. Gillman, Megan L. Grove, Nancy L. Heard-Costa, Scott R. Hutton, Rachel S. Kelly, Charles Kooperberg, Martin G. Larson, Jessica Lasky-Su, Deborah A. Meyers, Franklin P. Ockerman, Laura M. Raffield, Alexander P. Reiner, Stephen S. Rich, Jerome I. Rotter, Albert V. Smith, Kent D. Taylor, Ramachandran S. Vasan, Scott T. Weiss, Kari E. Wong, Alexis C. Wood, Prescott G. Woodruff, Lang Wu, Ronit I. Yarden, Junsun Yu, Laura Y. Zhou, Bing Yu, Degui Zhi, Han Chen

## Abstract

Modern large-scale genetic association analyses of imaging and omics data reveal unprecedented details of the genetic architecture of complex traits. Such analyses involve scanning thousands of phenotypes using linear mixed model-based genome-wide association study tools to control for sample relatedness. However, current LMM tools are not designed for such scale, creating a computational burden that hinders discovery. We propose TorchGWAS2, a cost-effective solution that overcomes the bottleneck using a deterministic variance-correction algorithm for LMMs, making it well-suited for GPU acceleration. TorchGWAS2 is applicable to unrelated and related individuals, cross-sectional and longitudinal studies, with and without missing data, and its computational complexity scales linearly with the number of phenotypes, genetic variants, and individuals. TorchGWAS2 showed more powerful association testing across 128 retinal image-derived endophenotypes of pairs of eyes from 64,703 UK Biobank participants and achieved two orders of magnitude speed-up analyzing 1,023 circulating metabolites in 16,352 Trans-Omics for Precision Medicine participants.

## Main

Large cohorts and studies have substantial data resources, and the volume of data continues to grow. They involve up to millions of samples, often with relatedness, and thousands of phenotypes, such as imaging-derived endophenotypes and omics measures. However, many currently available tools for scanning genetic associations for thousands of omics phenotypes, such as TensorQTL^1^ and TorchGWAS^2^, only implement linear regression models, which could lead to inflation of type I error rates when analyzing samples with family structures or samples with cryptic relatedness. In practice, related samples have to be excluded from the analysis, which reduces the effective sample size and results in a substantial loss of statistical power^3^.

Linear mixed models (LMMs) have been widely used in genome-wide association studies (GWAS) to account for sample relatedness. They model phenotypes with fixed covariates and genetic effects and a random intercept of which the covariance structure is proportional to a genetic relationship matrix (GRM) or a sparse kinship matrix. For longitudinal data, an additional random effect that changes over time for each individual can be included to account for within-individual variability from repeated measurements. Modern LMM-based GWAS tools often conduct score test^4, 5^, which means a model under the null of no fixed genetic effect is fitted to estimate variance components, polygenic background effect, and then the residuals of this null model are used to construct statistics for testing the association between phenotypes and genotypes. This is efficient for GWAS with millions of genetic variants because the mixed model, which is usually the computational bottleneck, is fitted only once. Researchers have invested significant effort in improving the computational efficiency of fitting mixed models and conducting association testing on large biobank-scale datasets. Methods such as BOLT-LMM^6^, fastGWA^7^, and REGENIE^8^ applied advanced algorithms to speed up the regression model fitting.

Furthermore, there is a widely used approach to speed up association tests by avoiding repeated large matrix computations, which applies a correction factor called GRAMMAR-Gamma^9^ (e.g. BOLT-LMM with infinitesimal model, SAIGE^10^, fastGWA) to reduce the computational complexity of testing each genetic variant from *O*(*N*^2^) to *O*(*N*), where *N* is the sample size. Almost all tools that use the correction factor estimate it by randomly selecting several null variants, which can be inefficient for a large number of phenotypes, as the program needs to repeatedly read large genotype files. For example, running BOLT-LMM, fastGWA, and REGENIE to test 50 quantitative traits with UK Biobank data across 30 million variants and 332,739 to 407,662 individuals required approximately 5.6 months of wall time, 401 central processing unit (CPU) days, and 126 CPU days, respectively, on cloud-based services Amazon Web Services (AWS) with 16 virtual CPU cores^8^. Despite the availability of large data resources and cloud computing, most research teams with limited computational resources cannot afford to analyze them. In addition, with a correction factor, the GWAS results depend on the random number seed used to sample the null variants, and the repeatability of is not always guaranteed. These limitations create a major computational bottleneck for large-scale studies involving thousands of phenotypes.

Additionally, existing GWAS tools are primarily designed for cross-sectional data; many large biobank studies increasingly collect longitudinal or repeated measurements for the same individuals. Extending scalable GWAS methods to these data presents additional computational challenges. To address these computational and technical bottlenecks, novel methods that compute corrections without reading genotypes are needed to advance GWAS of imaging and omics traits and to understand the genetic architecture of high-dimensional complex phenotypes, while equitizing data access for the broader scientific community. Therefore, we developed a new correction method that computes the correction factor deterministically without repeatedly accessing genotype files, which eliminates unnecessary I/O overhead. With such a correction factor, the step two regression can be readily accelerated by GPUs with implementations similar to existing tools such as TorchGWAS{Zhao, 2026 #106}. We packaged the two-step GWAS pipeline with this correction method and GPU-acceleration into a tool called TorchGWAS2.

TorchGWAS2 introduces a cost-effective and computationally scalable solution by jointly processing multiple phenotypes and using a deterministic variance correction factor for linear mixed models. In step 1, TorchGWAS2 fits a linear mixed model under the null of no fixed genetic effect, then computes a correction factor and applies it to the residuals. Then the corrected residuals are regressed on the genotype in a linear model in step 2, and another correction factor is applied to the standard errors. These correction factors are similar to those used in GRAMMAR-Gamma, BOLT-LMM, SAIGE, and fastGWA. But here we used a deterministic correction factor based on the expected value of genotype variation and covariance structure accounting for covariates and sample relatedness. The correction factors ensure the two-step approach remains statistically rigorous.

## Results

### Overview of TorchGWAS2

For a single-variant test, consider a linear mixed model

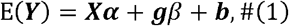

where ***Y***_*N*×1_ is the phenotype vector for *N* subjects, ***X*** is a *N* × *p* matrix of covariates including an intercept, and ***g***_*N*×1_is the genotype vector of a variant. ***α***_*p*×1_ is the fixed effect of covariates, *α* is the fixed genetic effect, and *b*~*N*(0, *λ***Ψ)** is the randomintercept for *N* subjects, where *λ* is the variance component parameter, and **Ψ** is an *N* × *N* GRM or a kinship matrix. Under the null hypothesis of no fixed genetic effect *H*_0_: *β* = 0, we fit the null model

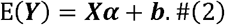

Denote residuals of the null model as ***r*** and scaled residuals as 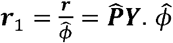 is the dispersion parameter estimate, and 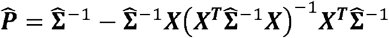 is the projection matrix, where 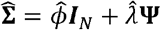 is the covariance matrix for samples estimated from the null model.

Let 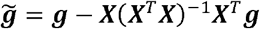 be the genotype vector adjusted by covariates, we define the correction factors

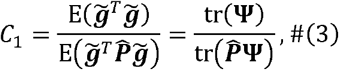

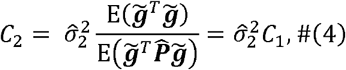

where 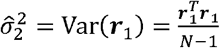 They play different roles through the pipeline. *C*_1_ is applied to ***r***_1_ in step 1 to generate the corrected residuals for unbiased effect estimation. *C*_2_ is applied to variance estimates in step 2 to correct for bias introduced by the linear model fitting. Specifically, in TorchGWAS2 step 2, we regress the corrected scaled residuals *C*_1_***r***_1_

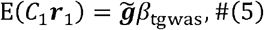

and the unbiased effect size estimate is

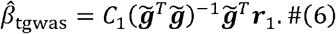

The corrected estimate of variance of 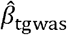 with *C*_2_ is

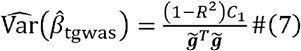

where *R* is the sample correlation between 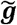 and ***r***_1_. The test statistic is

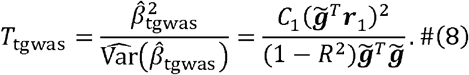

The correction factors are slightly modified for longitudinal data, as well as for missing phenotypes. Details are provided in Online Methods. We use a 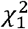 distribution to compute the TorchGWAS2 *P* value, which is a unified framework applicable to both cross-sectional and longitudinal data, with or without related individuals, with or without missing phenotypes.

Figure 1 shows the end-to-end data and file flowchart. After the linear mixed model is fitted and the correction factors are computed, TorchGWAS2 writes both the correction factors and the corrected residuals to an intermediate file. For samples with missing values, their residuals are assigned to zero and are excluded from contributing to the test statistic. These corrected residuals are then transferred to the graphics processing units (GPUs) along with the batched genotype data (Supplementary Figure S1). After computation, the GPU returns the regression outputs to the CPU, and TorchGWAS2 compiles them into a final output file containing the summary statistics for all phenotypes.

**Figure 1.**
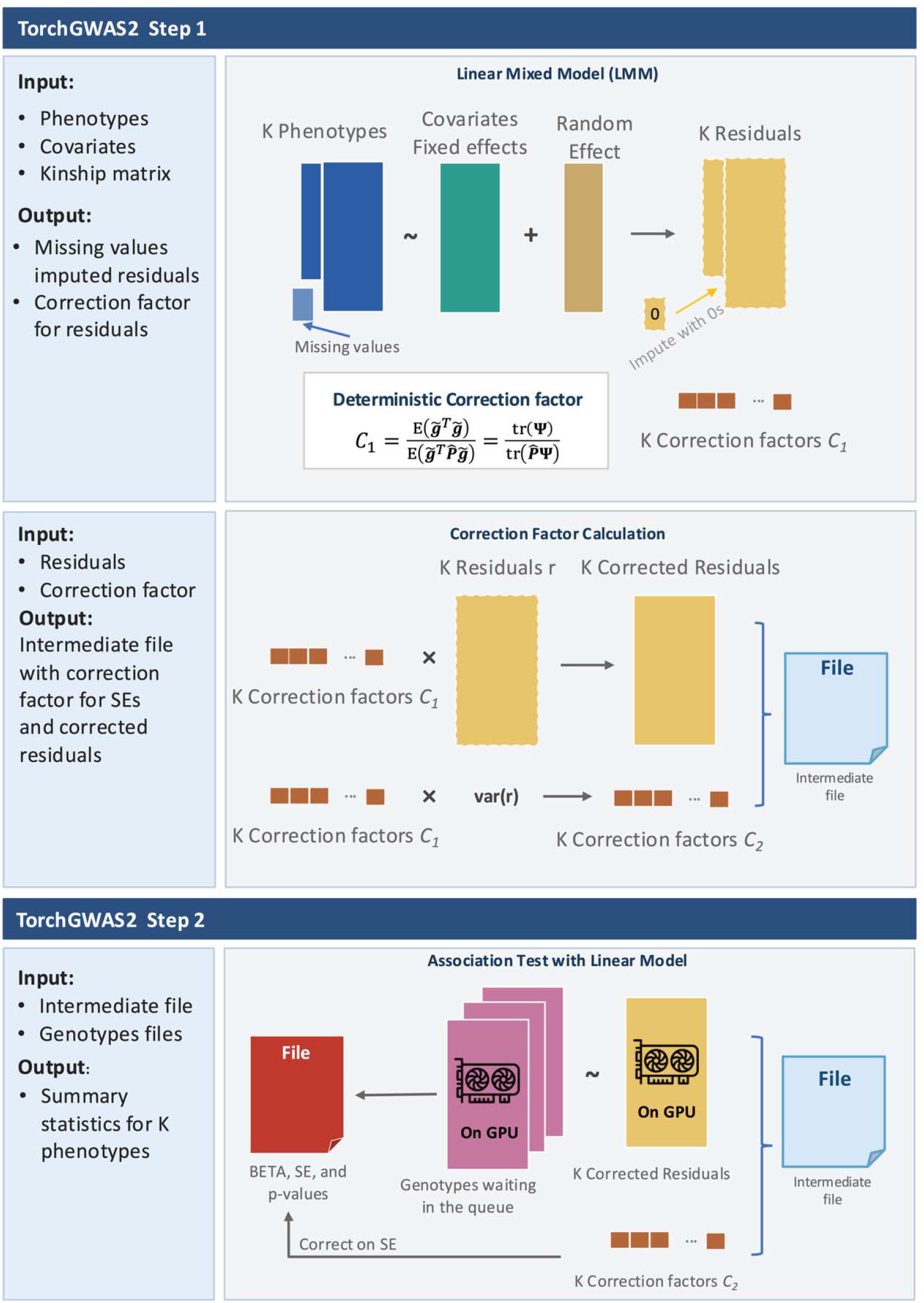
Workflow of TorchGWAS2 step 1 and step 2.

TorchGWAS2 accelerates its computation through the following five key optimizations. First, multiple phenotypes are analyzed in parallel so that genotype streaming and matrix operations are shared across traits. Second, the GRAMMAR-Gamma calibration factor for each phenotype is computed in closed form from mixed-model variance components and reused across all variants, without reading the genotype files. Third, the association test in TorchGWAS2 step 2 tests is performed using linear regression on residualized phenotypes and genotypes, thereby estimating the effect size of each variant as 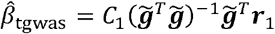 to avoid any *O*(*N*^2^) calculations. Fourth, for a streaming batch of *M* variants, *N* samples, and *P* phenotypes, ***r***_1_ for different phenotypes are processed jointly as a matrix rather than separately as individual vectors. This significantly reduces the I/O for reading genotype files, since each genotype block is read only once while being reused across all phenotypes. Finally, the matrix form of joint residuals and genotype blocks are highly efficient on modern hardware like GPUs, where the General Matrix Multiplication (GEMM) operations are implemented in parallel on the GPU and can exploit high floating-point throughput of GPUs.

### Simulation

We conducted comprehensive simulations to evaluate the performance of TorchGWAS2, including the empirical type I error rates, empirical power, runtime, and peak memory footprint across various scenarios. These results were then compared against fastGWA and REGENIE. We simulated thousands of replicates of 1 million common variants (MAF > 0.001) for 20,000 samples without relatedness and 100,000 samples with relatedness, which were sampled from a pool of 170,000 related samples consisting of extended and nuclear families. Both cross-sectional and longitudinal phenotypes with 5 repeated observations for each individual were generated from linear mixed-effect models. The details of data generation model can be found in the Online Methods. FastGWA and REGENIE were originally developed for cross-sectional data and their performance has not been verified in longitudinal data with repeated measures. In this setup, each repeated observation from the longitudinal data was treated as a distinct hypothetical individual sharing the same genome, thus the kinship matrix and genotypes were duplicated 5 times for the benchmark with fastGWA and REGENIE. Overall, we simulated four scenarios: cross-sectional and longitudinal analyses, each with unrelated and related samples. The detailed settings including the number of replicates, sample sizes, and genetic architecture are shown in Supplementary Table S1.

We compared the empirical type I error rate across all simulations at genome-wide significance level of 5 × 10 ^−8^ (Supplementary Figure S2 and Supplementary Table S2). This rate was calculated as the proportion of false positive findings out of the total number of null variants being tested. TorchGWAS2 presented well-controlled type I error rates across all scenarios with empirical rates close to the nominal threshold, while fastGWA and REGENIE showed inflated type I error rates when applied to longitudinal data with related samples. In cross-sectional data with related samples and longitudinal data with unrelated samples, fastGWA appropriately accounted for relatedness, while REGENIE was not well-calibrated, as expected behavior in the presence of high levels of relatedness as acknowledged by the authors^8^.

For the scenarios and methods with valid type I error control, we evaluated empirical power at 5 × 10^−8^ significance level, which is calculated as the proportion of true positive findings out of the total number of the causal variants being tested (Supplementary Table S2). All methods achieved comparable power in the simulation of complex traits GWAS with multiple independent associations, in scenarios when their results are valid (Supplementary Figure S2). In simulated quantitative trait loci (QTL) studies for molecular traits with one strong genetic association, TorchGWAS2 demonstrated increased power in cross-sectional data with related samples, and longitudinal data with unrelated samples.

We further evaluated the computational efficiency of three methods in terms of wall time and peak memory usage, using 20 parallel threads for all methods. In general, TorchGWAS2 running on a GPU (TorchGWAS2-GPU) showed a great wall time advantage when analyzing multiple phenotypes (Supplementary Table S3). In the cross-sectional related-sample scenario with 100,000 samples, TorchGWAS2-GPU completed 100 phenotypes in 306 seconds, compared to 7,977 seconds for fastGWA and 5,304 seconds for REGENIE, which showed speedups of 26-fold and 17-fold, respectively. A higher peak memory footprint was observed for TorchGWAS2, in which each thread had a default batch size of 1,000 variants in memory and waited in the queue for GPU consumption (Supplementary Figure S1). This memory footprint can be reduced by using a smaller batch size of 100 variants, which lowered peak memory usage at the cost of a moderate increase in the wall time (Supplementary Table S3).

### Application to UK Biobank retinal image data

To assess the ability to handle repeated measures within individuals, we applied TorchGWAS2 to UK Biobank retinal image data, which naturally have two measurements per individual. In previous work on retinal imaging GWAS^11^, analyses were performed separately for the left and right eyes with BOLT-LMM due to lack of computationally efficient tools to jointly analyze both eyes. TorchGWAS2 enables such joint analyses while accounting for the relatedness between left and right eyes. Therefore, we conducted GWAS jointly across both eyes and separately for the left and right eyes to evaluate computing time and memory footprint. We benchmarked TorchGWAS2 with two computationally efficient tools, REGENIE and fastGWA. The detailed study design is provided in the Online Methods.

TorchGWAS2 substantially reduced the runtime (Table 1). In single-eye analyses, TorchGWAS2-GPU was ~19 times faster than REGENIE and ~22 times faster than fastGWA. In both-eye analyses, TorchGWAS2 was 38.9 times faster than REGENIE and 48 times faster than fastGWA. This efficiency advantage can generalize to other types of repeated measurements and longitudinal phenotypes. TorchGWAS2-CPU, which utilizes the same algorithm as TorchGWAS2-GPU but runs association tests on CPUs, was still 2 to 5 times faster than REGENIE and fastGWA. TorchGWAS2 showed a larger peak memory footprint compared to REGENIE and fastGWA, with peak usage reaching up to 16 GB versus 11 to 12 GB for REGENIE and 1 to 3 GB for fastGWA (Supplementary Table S4). Although TorchGWAS2 requires more memory, this can be mitigated by adjusting the batch size of the genotypes, as demonstrated in the simulations (Supplementary Table S3).

**Table 1.** Benchmark of wall time in the application of UKB retinal imaging data for 128 dimensions.

| Benchmark | TorchGWAS2 |  |  | REGENIE |  |  | fastGWA |  |  |
| --- | --- | --- | --- | --- | --- | --- | --- | --- | --- |
|  | CPU runtime (sec) | GPU runtime (sec) | Speedup (GPU vs CPU)<sup>(1)</sup> | CPU runtime (sec) | CPU runs speedup | GPU runs speedup | CPU runtime (sec) | CPU runs speedup | GPU runs speedup |
| Both-eyes |  |  |  |  |  |  |  |  |  |
| Step 1 | 37 | 37 | – | 3248 | 87.8x | 87.8x | 619.5 | 16.7x | 16.7x |
| Step 2 | 1218 | 90 | 14.5x | 2113 | 1.7x | 23.5x | 2931.0 | 2.3x | 32.6x |
| Step X<sup>(2)</sup> | 10 | 10 | – | – | – | – | 1690.9 | – | – |
| Total wall time | 21.1 min | 2.3 min | 9.2x | 89.4 min | 3.9x | 38.9x | 110.5 min | 5.2x | 48.0x |
| Left-eye |  |  |  |  |  |  |  |  |  |
| Step 1 | 42 | 42 | – | 1818 | 43.3x | 43.3x | 185.1 | 4.4x | 4.4x |
| Step 2 | 1126 | 82 | 13.5x | 698 | 0.6x | 8.5x | 1765.2 | 1.6x | 21.5x |
| Step X | 10 | 10 | – | – | – | – | 169.0<sup>(3)</sup> | – | – |
| Total wall time | 19.6 min | 2.2 min | 8.9x | 42.0 min | 2.1x | 19.1x | 48.7 min | 2.5x | 22.1x |
| Right-eye |  |  |  |  |  |  |  |  |  |
| Step 1 | 25 | 25 | – | 1788 | 71.5x | 71.5x | 193.2 | 7.7x | 7.7x |
| Step 2 | 1192 | 91 | 11.1x | 736 | 0.6x | 8.1x | 1874.4 | 1.6x | 20.6x |
| Step X | 21 | 21 | – | – | – | – | 94.0<sup>(4)</sup> | – | – |
| Total wall time | 20.6 min | 2.3 min | 6.4x | 42.1 min | 2.0x | 18.3x | 50.1 min | 2.4x | 21.8x |

The scatter plots show high consistency between TorchGWAS2 and fastGWA results in both single-eye analyses (Supplementary Figure S3) and both-eye analyses (Figure 2). REGENIE results for single-eye analyses were also consistent with TorchGWAS2 and fastGWA, but underestimated standard errors and inflated results were observed in REGENIE both-eye analyses (Figure 3, genomic inflation factor ranging from 1.144 to 1.443 with a mean of 1.254), due to high levels of relatedness between left and right eyes.

**Figure 2.**
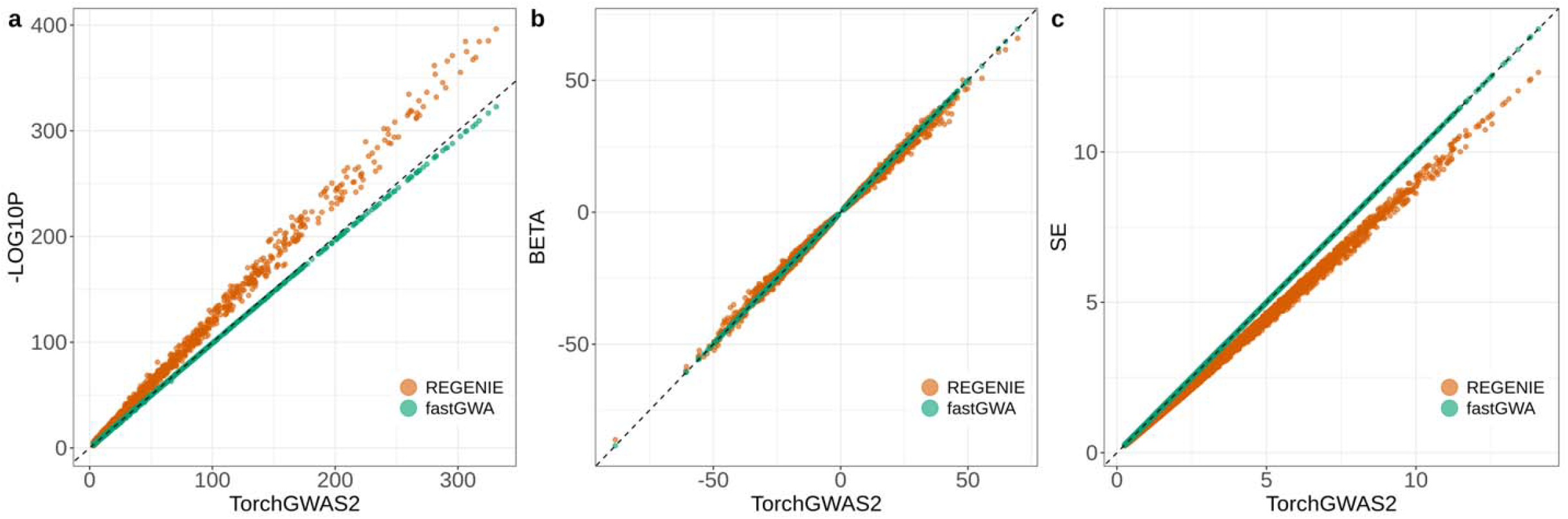
Comparison of *P* values on the −log_10_ scale (**a**), effect size estimates (**b**), and standard errors (**c**) among TorchGWAS2, fastGWA and REGENIE for both-eye analysis combined for all 128 dimensions from UKB retinal image data. The variants in the plots were filtered with MAF > 0.001, and *P* < 0.001. The x-axis represents results from TorchGWAS2 and y-axis represents results from fastGWA (green) and REGENIE (orange). The dashed black line represents the identity line.

**Figure 3.**
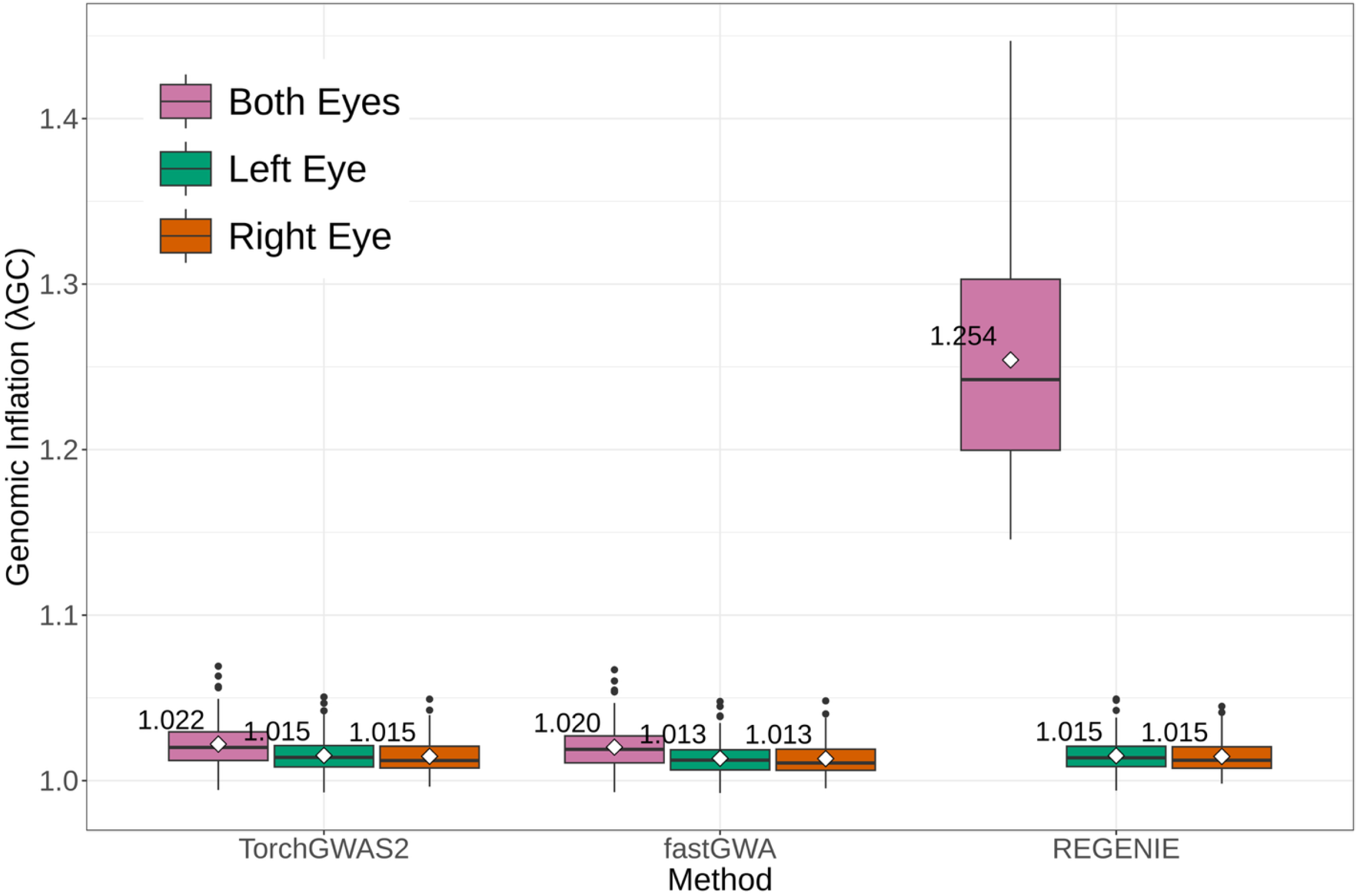
Comparison of genomic inflation factor (based on the median χ2 statistic) from UKB retinal image data across three methods (TorchGWAS2, fastGWA and REGENIE) and eyes in the analysis (both-eye, left-eye and right-eye). The white squares and the figures beside them are the means of the genomic inflation factors across 128 dimensions.

The Miami plot (Figure 4) compares both-eye and single-eye analyses with TorchGWAS2. The signals of both-eyes analyses were consistently stronger than single-eye analyses, which indicates that joint modelling of the left and right eyes leveraged information from both eyes and strengthened the association signals. Three genome-wide significant regions containing genes *GABRB1, FAM234B*, and *MTUS2* implicated in retinal neuron functions, were identified exclusively from both-eye analyses at the Bonferroni-corrected significance level of 5 × 10^−8^/128 = 3.9 × 10^−10^ for 128 endophenotypes.

**Figure 4.**
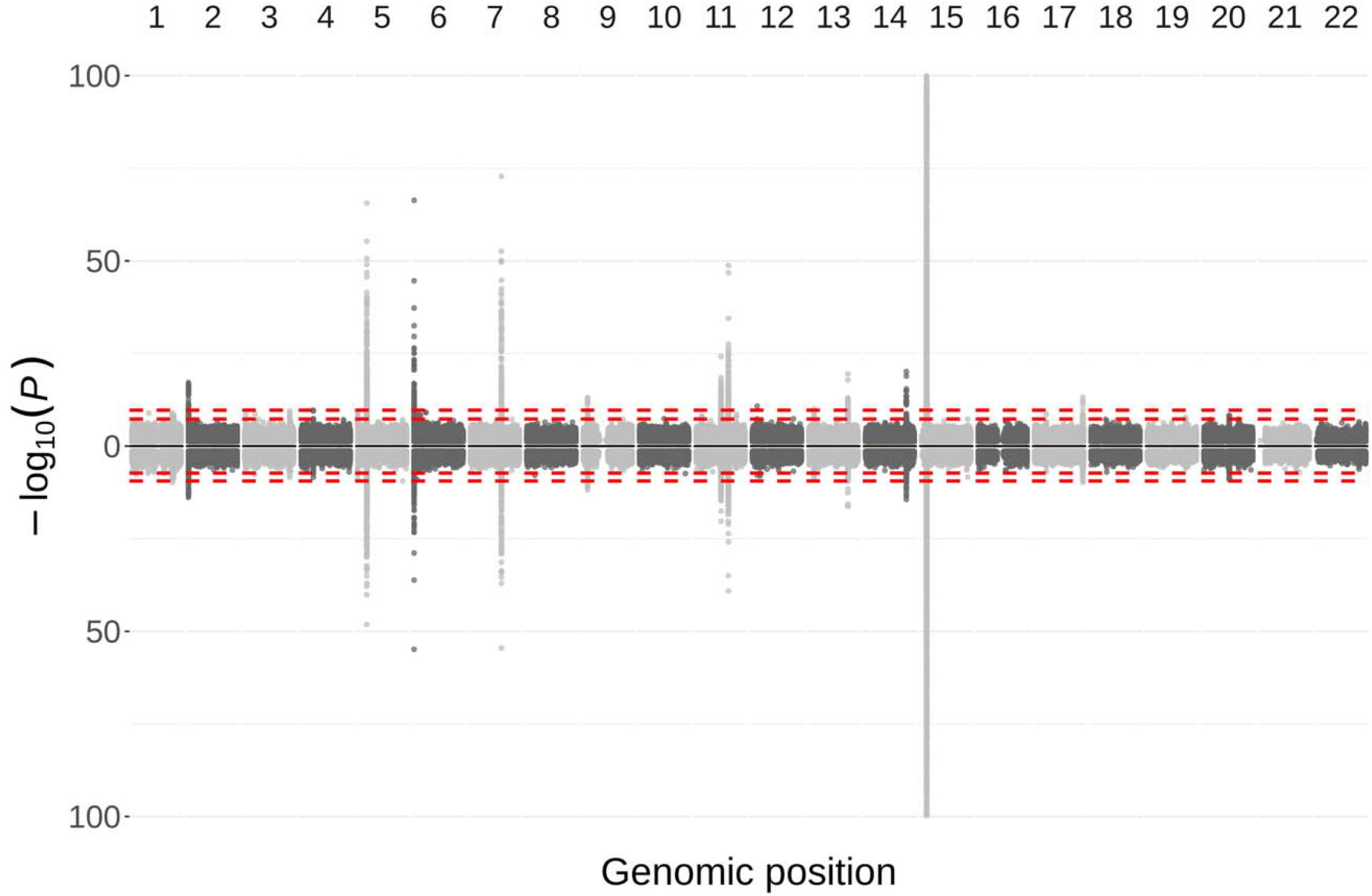
Miami plot of both-eye analysis (top) and single-eye analysis (bottom) combined for all 128 dimensions TorchGWAS2 *P* values from UKB retinal image data. Minimum *P* value between the left- and right-eye analyses was selected as the single-eye analysis *P* value for each variant. The dashed lines are −log_10_ (5 × 10^−8^) and −log_10_ (5 ×10 ^−8^/128) for both-eye analyses, and −log_10_ (5 × 10^−8^) and −log_10_ (5 × 10^−8^/256) for single-eye analyses, respectively. Only variants with MAF > 0.001 and *P* < 0.05 are plotted. The plot is cut off at −log_10_ (*P*) = 100 for better visualization. The significant regions from TorchGWAS2 both-eye analysis are provided in Supplementary Table S5.

Additionally, TorchGWAS2 was compared with the BOLT-LMM with the infinitesimal model (BOLT-LMM-inf) from previous work^11^ in single-eye analyses (Supplementary Figure S4), which showed high consistency between the two methods. We further generated a Miami plot (Supplementary Figure S5) comparing both-eye analyses performed with TorchGWAS2 and single-eye analyses performed with BOLT-LMM-inf, indicating that TorchGWAS2 both-eye analyses produced smaller *P* values at all loci (Supplementary Table S5).

### Application to TOPMed metabolomics data

To assess the performance of TorchGWAS2 on multi-ancestry samples with relatedness, we applied it to Trans-Omics for Precision Medicine (TOPMed) metabolomics data in QTL (metQTL) studies. In previous work^12^, genome-wide association analyses of these metabolomics data were conducted on the NHLBI BioData Catalyst platform using GMMAT^5^. However, due to complex family structure and sample relatedness, GMMAT required substantial computational resources, taking approximately 4 months of wall time with 96 threads in parallel and costing over $8,000. To evaluate TorchGWAS2 with TOPMed metabolomics data, metabolites were grouped into batches based on the study membership of the samples, and batches with fewer than 10 metabolites were excluded, resulting in 1,023 metabolites grouped into 11 batches. Details of the study design and data description are provided in the Online Methods.

We benchmarked TorchGWAS2 against REGENIE, fastGWA and GMMAT by evaluating genomic inflation factors and consistency of summary statistics on two typical batches: batch 6, which includes samples from the Framingham Heart Study (FHS)^13^ with family structures spanning three generations, and batch 9, which consists mostly of unrelated samples. The scatter plots (Supplementary Figure S6) showed high consistency between TorchGWAS2 and GMMAT. TorchGWAS2 yielded stronger signals for variants with large effects. For 1-Methylxanthine (HMDB0010738) in batch 9, the previously reported significant association with variant rs4921915 identified by GMMAT (*β* = 0.308 for the A allele as compared to the reference G allele, *P* = 2.60 × 10^−71^)^12^ was replicated by TorchGWAS2 with the same effect size estimate *β* = 0.308 and a smaller *P* value (*P* = 5.81 × 10^−74^). Comparisons between batch 6 and batch 9 showed that the discrepancy between TorchGWAS2 and REGENIE was amplified when substantial family samples from FHS were present. Among TorchGWAS2, GMMAT, and fastGWA, we found that fastGWA produced smaller median test statistics (Supplementary Figure S7) and larger *P* values for variants with large effects. This resulted in more conservative *P* values in the tail and is consistent with a previous UK Biobank application in 407,746 individuals of white British ancestry^8^.

We compared the runtime and costs of GPU-accelerated TorchGWAS2 runs on the Google Cloud computing environment via BioData Catalyst with GMMAT for all 11 batches (Table 2). TorchGWAS2-GPU completed all 11 batches analyses in 19.35 hours of runtime, which is 159-fold faster than months of total runtime (3,076 hours) for GMMAT running with 96 threads in parallel. Using the platform’s recorded billing charges, the total computation cost was $68 with TorchGWAS2-GPU, which is 127-fold cheaper than $8,664 with GMMAT.

**Table 2.** Comparison of runtime and costs between TorchGWAS2 (GPU) and GMMAT in the application of TOPMed metabolites data.

| Batch | Sample size | No. metabolites | GMMAT time (hr) | TorchGWAS2 time (hr) | Speedup | GMMAT cost | TorchGWAS2 cost | Cost savings |
| --- | --- | --- | --- | --- | --- | --- | --- | --- |
| 1 | 2,469 | 44 | 95.96 | 1.33 | 72.1x | \$283.14 | \$9.70 | 29.2x |
| 2 | 4,045 | 11 | 35.23 | 1.28 | 27.6x | \$78.74 | \$5.49 | 14.3x |
| 3 | 5,043 | 14 | 45.39 | 1.29 | 35.1x | \$132.83 | \$4.60 | 28.9x |
| 4 | 2,778 | 73 | 169.72 | 1.45 | 116.9x | \$348.39 | \$6.47 | 53.8x |
| 5 | 5,799 | 47 | 184.69 | 1.57 | 117.9x | \$452.08 | \$5.23 | 86.4x |
| 6 | 6,797 | 124 | 485.32 | 2.05 | 236.6x | \$1364.89 | \$5.24 | 260.5x |
| 7 | 6,844 | 21 | 42.68 | 1.44 | 29.6x | \$141.63 | \$5.26 | 26.9x |
| 8 | 8,223 | 17 | 34.86 | 1.34 | 26.0x | \$128.50 | \$4.89 | 26.3x |
| 9 | 8,768 | 433 | 896.47 | 2.70 | 331.8x | \$3329.21 | \$7.27 | 457.9x |
| 10 | 14,567 | 93 | 411.03 | 2.18 | 188.2x | \$881.04 | \$6.83 | 129.0x |
| 11 | 15,565 | 146 | 674.40 | 2.71 | 249.1x | \$1523.90 | \$7.06 | 215.8x |
| Total | — | 1,023 | 3075.75 | 19.35 | 159.0x | \$8664.35 | \$68.04 | 127.3x |

Notes
(1) Costs were charged based on the total running time, which includes the time for uploading input files to the cloud, the execution of TorchGWAS2, and downloading the results.

## Discussion

To our knowledge, TorchGWAS2 is the first tool to integrate GPU acceleration with a deterministic variance correction factor to facilitate large-scale GWAS using linear mixed models. The correction factor has been widely used to reduce the computational complexity of testing each genotype from *O*(*N*^2^) to *O*(*N*) in linear mixed models. Existing methods, such as GRAMMAR-Gamma^9^, BOLT-LMM^6^, SAIGE^10^, and fastGWA^7^ rely on randomly sampling a subset of variants to estimate correction factors and therefore incur heavy I/O overhead for analyzing thousands of phenotypes. In contrast, TorchGWAS2 computes the correction factor analytically using a deterministic algorithm that does not depend on the random number seed, eliminating sampling noise and enabling scalable, accurate, and reproducible analyses. TorchGWAS2 also accommodates missing values in phenotypes, allowing analyses with incomplete observations. This represents a major advance in high-dimensional GWAS methodology by combining GPU acceleration with a deterministic algorithm.

TorchGWAS2 showed consistent results with existing tools, and even some modest improvements in statistical power. First, it increases the power of the association test and maintains calibrated type I error. Under the assumption of an infinitesimal model in which a trait is influenced by a very large number of variants, each with a very small effect size^14^, people have been using mixed-effect models and the score test to conduct GWAS among related samples. The score test estimates the residual variance and heritability under the null, so they are fixed across variants, but they can be overestimated if the variant effect size is large, leading to overestimated standard errors and reduced statistical power. In TorchGWAS2, the residual variance estimate is updated for each variant based on its correlation with residuals as shown in Equation (7); thus, standard errors are smaller than score test-based approaches such as GMMAT^5^ and fastGWA^7^ for a large-effect variant. This feature of TorchGWAS2 was demonstrated in the simulation of QTL studies (Supplementary Table S2). In the presence of relatedness and a single strong signal, TorchGWAS2 shows greater power compared to fastGWA (cross-sectional data with related samples: 0.45 vs. 0.36, longitudinal data with unrelated samples: 0.71 vs. 0.65). Additionally, in cross-sectional data with unrelated samples, both TorchGWAS2 and fastGWA fit a linear model with the Wald test which used different residual variance for different variants, resulting in comparable power between the two methods in QTL studies (0.66 vs. 0.65).

Powerful association test performance for TorchGWAS2 is further supported by its application to the UK Biobank retinal image data and TOPMed metabolomics data (Figure 2, Supplementary Figure S3, Supplementary Figure S6). In the UK Biobank retinal image data, we identified three genome-wide significant regions containing genes *GABRB1, FAM234B*, and *MTUS2* from both-eye analyses (Supplementary Table S5). These same regions were also identified by fastGWA (TorchGWAS2 vs. fastGWA: rs9838604, 2.79E-10 vs. 2.85E-10; rs4694845, 1.96E-10 vs. 2.02E-10; rs1684387, 1.59E-11 vs. 1.61E-11; rs11616792, 8.73E-11 vs. 8.29E-11). REGENIE was not included in this discussion since its inflation in simulation with repeated measurements scenario. *GABRB1* encodes beta-1 subunit of the GABA type A receptor, which is essential for inhibitory neurotransmission. GABAergic inhibition plays a critical role in retinal visual processing^15^. *FAM234B* shows detectable mRNA expression in human retinal tissue (The Human Protein Atlas^16^). *MTUS2* encodes a microtubule plus-end tracking protein that regulates the structural scaffold that supports intracellular transport in neurons^17^, and shows enhanced expression in retinal amacrine cells, retinal bipolar cells and retinal ganglion cells (The Human Protein Atlas^16^). These discoveries were previously missed in single-eye analyses^11^, illustrating the advantage of leveraging multiple repeated measures from the same individuals, an analytical strategy empowered by TorchGWAS2.

In the metQTL studies, the scatter plot of − log_10_ versus effect sizes (Supplementary Figure S8) shows that variants at the tail are mostly with larger effect sizes. For those variants, TorchGWAS2 produced smaller standard errors and lower *P* values while effect size estimates were similar to those of other methods. As an example, we looked at the lead variant for 1-Methylxanthine in batch 9, in which most of the samples came from COPDGene and SPIROMICS studies. It is a metabolic byproduct of caffeine. Smoking induces the liver enzyme Cytochrome P450 1A2 (CYP1A2), which accelerates caffeine clearance and subsequent formation of downstream metabolites, including 1-Methylxanthine^18^, therefore leads to its higher blood concentration among current smokers^19^.

Second, TorchGWAS2 directly models within-subject variance, whereas other existing tools assume one observation per individual and need to be adapted to handle repeated measures or longitudinal data. For data with repeated measurements, the sample identifiers are usually repeated in the phenotype or covariates file. For example, in the UK Biobank retinal image data, the left eye and right eye are repeated observations from the same person. FastGWA and REGENIE were not originally developed for repeated measures in the input file^7, 8^. While duplicating the GRM and individual genotypes allowed fastGWA to analyze such data as hypothetical monozygotic twins with well-controlled type I error rates, REGENIE was not designed to handle such high levels of relatedness. Moreover, duplicating the genotype files resulted in doubled I/O for both fastGWA and REGENIE. Notably, while fastGWA appropriately accounts for related samples in cross-sectional data or repeated measures from unrelated samples in longitudinal data, it utilizes a single variance component for random effects, which may cause convergence issues during null model fitting and inflated type I error rates for repeated measures from related samples in longitudinal data (Supplementary Figure S2d and Supplementary Table S2). This highlights the advantage of TorchGWAS2 in analyzing repeated measures in longitudinal data, for both unrelated and related samples, as it directly models within-subject and between-subject variance components while controlling the type I error rates and maintaining computational efficiency.

Third, TorchGWAS2 conducts an available-case analysis but handles phenotypes that are missing completely at random efficiently. In large biobank studies, many traits are measured across different subsets of individuals, leading to varying missing rates across phenotypes. Unlike tools such as TensorQTL^1^, TorchGWAS2 allows keeping samples with missing phenotypes in the analysis without requiring any imputation prior to analysis.

Fourth, TorchGWAS2 is cost-effective when running large-scale phenome- and genome-wide scans on cloud-based computation platforms. Many major genomic research platforms are powered by cloud infrastructure such as Amazon Web Services (AWS) and Google Cloud, including NHLBI BioData Catalyst, the UK Biobank Research Analysis Platform (RAP), the NCI Cancer Research Data Commons (CRDC), and the NIH *All of Us* Researcher Workbench. These platforms charge users based on the specifications and runtime on the computation nodes. TorchGWAS2 significantly reduces runtime, making it highly suitable for cloud environments. As a result, TorchGWAS2 allows researchers to conduct comprehensive GWAS faster and more cost-effectively, and lowers the computational barrier to large-scale genetic discovery.

In summary, TorchGWAS2 is a GPU-accelerated software tool designed to support phenome- and genome-wide association studies in related samples. It is the first to incorporate a deterministic variance correction factor for linear mixed models, eliminating sampling variability in existing approaches with correction factors estimated using randomized algorithms, and enabling scalable, accurate, and fully reproducible analyses. By conducting more powerful association tests and reducing the runtime, TorchGWAS2 represents a significant advance in high-dimensional GWAS methodology. As biobank studies increasingly collect longitudinal and multi-modal phenotypes (e.g. *All of Us*, UK Biobank, and others), fast mixed-model tools that can account for both related samples and longitudinal data are needed to further advance the field, especially for imaging and omics genetics. TorchGWAS2 reduces computational costs, shortens analysis time from months to hours, and makes large-scale multi-phenotype GWAS feasible for investigators with limited computational resources.

## Data Availability

All data produced in the present study are available upon reasonable request to the authors.

## Code and data availability

TorchGWAS2 is available at https://github.com/hanchenlab/TorchGWAS2/tree/support-to-bed-format. The individual-level UK Biobank data are available to qualified researchers through the UK Biobank Access Management System upon application approval and in accordance with the UK Biobank’s data access policies. The individual-level TOPMed whole-genome sequencing and phenotype data analyzed in this study are available through controlled-access NIH repositories (e.g. dbGaP) to qualified investigators who obtain the appropriate approvals. Data sharing and access to TOPMed data are conducted in accordance with NIH Data Sharing and Management policies.

## Acknowledgement

This work was supported by NIH grants R01 EY032768, U01 AG070112 and R01 AG081398. This research has been conducted using the UK Biobank Resource under application number 24247. Molecular data for the TOPMed Program was supported by the National Heart, Lung, and Blood Institute (NHLBI). See the Cohort Description section in the supplemental information for study-specific omics support information. CORE support including centralized genomic read mapping and genotype calling, along with variant quality metrics and filtering were provided by the TOPMed Informatics Research Center (3R01HL-117626-02S1; contract HHSN268201800002I). CORE support including phenotype harmonization, data management, sample-identity QC, and general program coordination was provided by the TOPMed Data Coordinating Center (R01HL-120393; U01HL-120393; contract HHSN268201800001I). We gratefully acknowledge the studies and participants who provided biological samples and data for TOPMed. L.W. is supported by U54HG013243. Additional study-specific acknowledgments are included in supplemental information.

## Disclaimers

Dr. Yarden previously had an NHLBI programmatic role with one co-author on matters unrelated to this manuscript, and no associated funding supported this work. The views expressed in this manuscript are those of the authors and do not necessarily represent the views of the National Heart, Lung, and Blood Institute; the National Institutes of Health; or the U.S. Department of Health and Human Services.

## Online Methods

### Linear mixed model and the score test

For a single-variant test, consider a linear mixed model

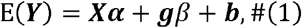

where ***Y***_*N*×1_ is the phenotype vector for *N* subjects, ***X*** is a *N* × *p* matrix of covariates including an intercept, and ***g***_*N*×1_ is the genotype vector of a variant. ***α***_*N*×1_ is fixed effect of covariates, *β* is the fixed genetic effect, and *b*~*N*(0, *λ***Ψ)** is random intercept for *N* subjects, where *λ* is the variance component parameter, and **Ψ** is a *N* × *N* genetic relation matrix (GRM) or 2× kinship matrix. Under the null hypothesis of no fixed genetic effect *H*_0_: *β* = 0, we fit the null model

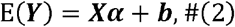

and get the residuals *r*, and scaled residuals 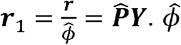 is the dispersion parameter estimate, and 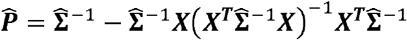 is the projection matrix, where 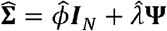 is the covariance matrix for samples estimated from the null model.

For longitudinal data, denote *N*_*obs*_ as the number of observations for *N* subjects, and the residual vector 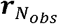 with length *N*_*obs*_ is compressed to the residual vector ***r*** with length *N* by 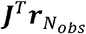, where 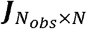 is the mapping matrix from observations to subjects. Specifically for observation *i* that is measured in subject *j*, ***J***_*i,j*_ = 1. The variance matrix is 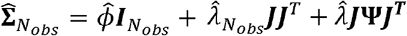, where 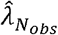 is the variance component parameter estimate for non-genetic random individual effect to account for within-individual relatedness, and 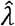 is variance component parameter estimate for polygenic random individual effect attributed to **Ψ**. The projection matrix is 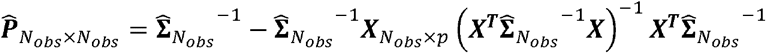.

A widely used score test for the linear mixed model in GWAS is constructed with the score ***U*** = *g*^T^***r***_1_, and the variance of the score is 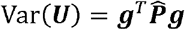. Therefore, the score test statistic is

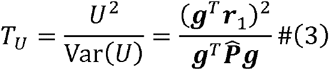

where *T*_*U*_ follows a 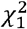 distribution. If the variant effect is too small to change the residual variance estimate, the variant effect estimate can be approximated by

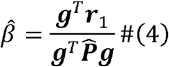

with a variance estimate of

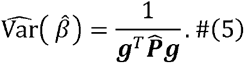

### Bias in the variance estimate and its correction

Denoting 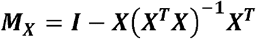, and 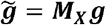, a linear model regresses the scaled residuals ***r***_1_ on genotypes with

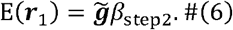

where 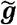 is the genotype vector after adjustment for covariates, and it is centered. The corresponding effect estimate

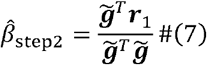

and the variance of the estimate is

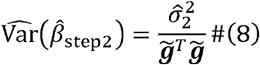

where 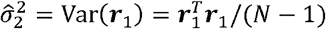. They are biased in the presence of related individuals or longitudinal data, as the variance estimate fails to account for sample relatedness.

However, the computation of 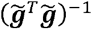 in the linear model is more efficient than the computation of 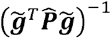, with computation complexity of *O*(*N*) versus *O*(*N*^2^). Therefore, researchers have taken advantage of this efficiency by using a correction factor called “GRAMMAR-Gamma” to approximate the variance^6, 7, 9, 10^. The “GRAMMAR-Gamma” is defined as 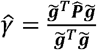, and the corrected variance of variant effect is 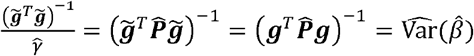. Assuming that the effect of each variant on the trait is small, the correction factor for each variant is similar^9^. With this assumption, the genome-wide estimate of 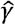 is usually approximated by the mean of 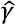 from randomly selected 20 to 1000 null variants. This process usually requires additional reading of the genotype file before the association test, which incurs additional I/O, and the correction factor estimate depends on the random number seed. To address this computation inefficiency, we derived two deterministic correction factors.

### Derivation of TorchGWAS2 deterministic correction factors

Assuming non-inbreeding individuals from multiple ancestries with 2 kinship matrix or a GRM and ancestry principal components (PCs) are included in the covariates, we define a working variance of 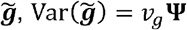 and 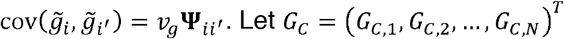 be the genotype count vector for single variant across *N* individuals, wher *G*_*C,i*_ ∈ {0,1,2} denotes the number of copies of the effect allele carried by individual *i*. We define the scala *v*_*g*_ = *var*(*G*|*X*) the genotype variance conditional on covariates. Therefore, genotypes can be written a 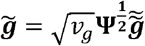, wher 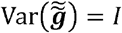. Define the correction factors

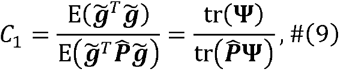

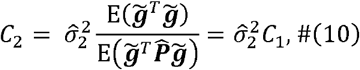

Where 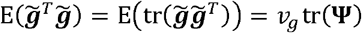, and 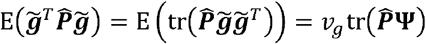. The quantity 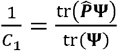 is similar to 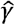 defined above. While 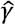 is estimated by averaging across several random variants, 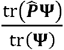 is deterministic and determined by the relatedness matrix **Ψ**. Under the same assumption of estimating 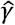, we approximate 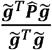 by a constant across all variants, which is 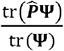. For longitudinal data, 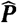 is a *N*_*obs*_ × *N*_*obs*_ matrix, and the correction factors are

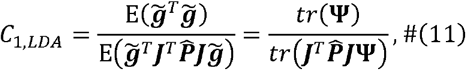

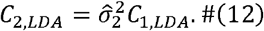

They play different roles through the pipeline. *C*_1_ is applied to ***r***_1_in step 1 to generate the corrected residuals for unbiased effect estimation. *C*_2_ is applied to variance estimates in step 2 to correct for bias introduced by the linear model fitting. Specifically, in TorchGWAS2 step 2, we regress the corrected scaled residuals *C*_1_ ***r***_1_

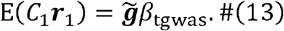

And the effect size estimate is

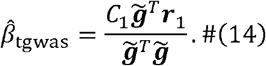

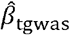 is unbiased since

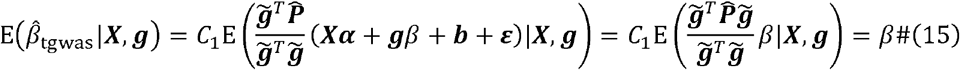

With 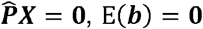, and expectation of random error E(***ε***) = **0**. Denote the residual variance of model (13) a 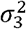. The 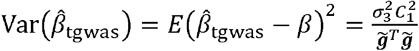, and the estimate of 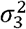 is 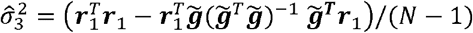. Therefore, the corrected estimate of variance of 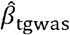 with *C*_2 is_

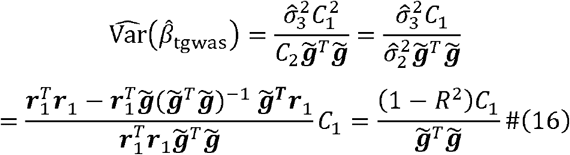

where and *R* is the sample correlation coefficient between 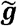 and ***r***_1_. The expectation of variance estimator is

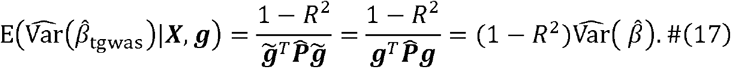

The test statistic is

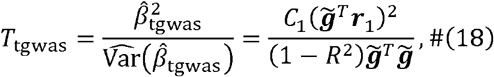

which asymptotically follows 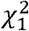. When a variant is correlated with the scaled residuals ***r***_1_, the TorchGWAS2 standard errors of its genetic effect would be smaller than the standard errors estimated under the null hypothesis of no genetic association, increasing the power of TorchGWAS2 compared to the score test.

For unrelated samples, the framework of TorchGWAS2 is closely related to the partial F test in linear regressions comparing the null model with residual variance 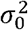 and the full model E(***Y***) = ***Xα*** + ***g****β* with residual variance 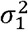. For partial F test, residualizing both phenotype and covariates gives ***r***_*Y*_ = ***M***_***X***_***Y*** and 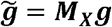 Residual sum of squares under the null model is RSS_0_ = ***Y***^**T**^***M***_***X***_***Y*** and residual sum of squares under the full model is 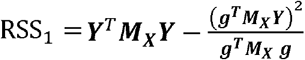. Therefore, the estimated residual variance for the null model and the full model are 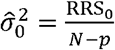 and 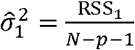, The partial F test for *H*_0_: *β* = 0 is

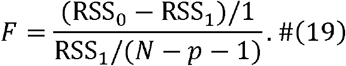

By Frisch–Waugh–Lovell theorem, the estimate of variant effect size from the full model is identical to the estimate from the residualized model 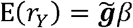

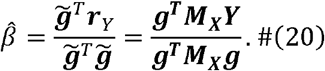

The variance estimator is

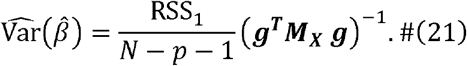

For TorchGWAS2 without related samples, 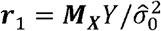 and 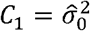. The estimate of effect size is

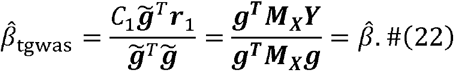

The variance estimate is

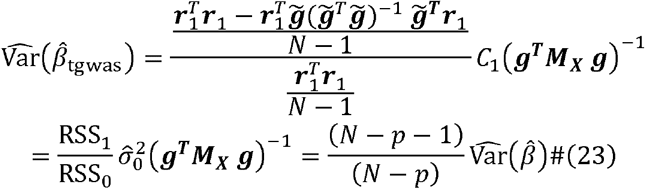

The test statistic is

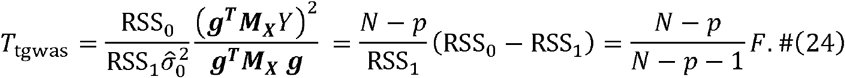

In the linear regression, *P* value from the partial F test is calculated using an F distribution with 1 and *N* − *p* − 1 degrees of freedom, whereas in TorchGWAS2 we use a chi-square distribution with 1 degree of freedom to better accommodate compatibility with related samples or longitudinal data. In large unrelated samples of cross-sectional data, TorchGWAS2 and F test statistics and *P* values would be very close.

### Handling missing phenotypes

TorchGWAS2 can handle missing phenotypes using an available-case analysis while maintaining a rectangular data structure for efficient computation. Specifically, individuals with missing phenotypes are not included when fitting the null model, but their residuals are set to zeros to make sure the residual vectors of all phenotypes have the same length and can be stacked to a matrix and processed simultaneously in step 2 GWAS. This procedure is not an imputation strategy to handle missing data, but it avoids taking a subset of samples for each phenotype repeatedly in step 2 GWAS and could leverage this rectangular matrix for GEMMs on GPU. The validity of this approach relies on the assumption that phenotypes are missing completely at random (MCAR).

Denote a *N* × 1 phenotype vector 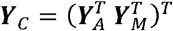 with *N*_*A*_ available phenotype values *Y*_*A*_ and *N*_*M*_ missing values *Y*_*M*_, and *N* = *N*_*A*_ + *N*_*M*_. TorchGWAS2 fits the linear mixed null model with available data and computes the scaled residuals of available phenotype ***r***_1*A*_. Correction factor *C*_1_ is computed as

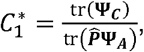

where the kinship matrix or GRM **Ψ**_*A*_ only contains individuals that are available for this phenotype, and ***Ψ***_*C*_ includes all individuals in complete data. ***r***_1*A*_ is expanded to 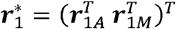 with residuals for missing data ***r***_1*M*_ = **0**. Correction factor *C*_2_ is computed as

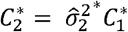

where 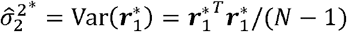.

### Simulations

Denote *i* = 1, …, *N* as individual index, *t* = 1, …, *T* as time points index, with *T* = 1 for longitudinal data and *T* = 1 for cross-sectional data. The covariates of fixed effect, including age, sex, and smoking status are the same across all scenarios. Age for individual *i* A at time *t* is age_*it*_ = age_*i*1_ + 5 (t − 1), where the baseline age_*i*1_ ~ Uniform(20,60). Smoking status smk_*it*_ ~ Bernoulli(0.3). We considered the following data generation models.

Model 1 Longitudinal null model with *T* = 5 for each individual

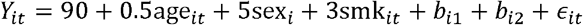

Model 2 Longitudinal model with genetic association with *T* = 5 for each individual

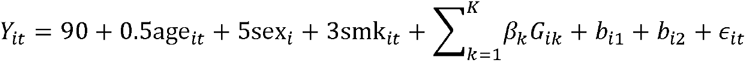

where *β*_*k*_ is genetic effect of causal variant *k, G*_*ik*_ is genotype of individual *i* with MAF range from 0.05 to 0.2, *b*_*i*1_ is an element in vector *b*_1_ ~ *N*(0,4**Ψ**) which is polygenic random effect capturing familial genetic correlation via the kinship matrix **Ψ**, *b*_*i*2_ is an element in vector *b*_2_ ~ N (0,2*I*_*N* × *N*_) which is the random individual effect not attributed to the relatedness from **Ψ** both *b*_*i*1_ and *b*_*i*2_ are the same across *t*, and *ϵ*_*it*_-~ (0,5) is the random error. The cross-sectional data is obtained from the baseline of the longitudinal data, and the missing rate of the phenotypes ranges from 0% to 30%.

Family structure was simulated with multiple steps. A total of 170,000 samples were simulated from 4,000 extended families with three generations and 25,500 quartet families. Each extended family consisted of 2 founders in the first generation, 3 offspring and 3 external partners in the second generation, and 9 offspring in the third generation. The pool of unrelated samples was defined as 5 founders per extended family and 2 founders per quartet family. From 170,000 simulated samples, 80,000 related samples were sampled from the non-founder individuals, and 20,000 unrelated samples were sampled from the founder pool. These 100,000 samples were combined into the full sample set containing both related and unrelated individuals.

For empirical type I error rates computation, 1900 replicates of phenotypes were simulated for 100,000 samples with relatedness and 20,000 without relatedness. We computed the empirical power in two scenarios, 1) GWAS of complex traits with *K* = 50 independent causal variants, and together they contribute to 2% (unrelated samples) or 0.5% (related samples) phenotypic variance in cross-sectional data and 1% (unrelated samples) or 0.2% (related samples) phenotypic variance in longitudinal data; 2) QTL studies with 400 samples and only one causal variant which contribute to 2% phenotypic variance in cross-sectional data and 1% phenotypic variance in longitudinal data. Each scenario has 100 replicates. Details of simulation settings can be found in Supplementary Table S1.

### UK Biobank retinal image data

In UK Biobank dataset, we restricted the analyses to 64,703 self-reported white British (field: 21000) and genetically clustered as Caucasian (field: 22006). Therefore, there are 129,406 images including both left- and right-eye fundus images (field: 21015 and 21016). For each image, 128 self-supervised image-derived phenotypes were extracted from UK Biobank eye image data from the pretrained model^11^. We analyzed 658,720 directly genotyped single nucleotide polymorphisms (SNPs) from the UK Biobank Axiom (field: 22438) with minor allele frequency (MAF) > 0.001. Covariates age, sex, PC1-PC10 were included in the model. Related individuals were included, and sample relatedness was accounted for using a sparse kinship matrix (field: 22012) in the linear mixed model.

GWAS was conducted (1) separately for left eye and right eye and (2) jointly for left and right eye. We benchmarked the performance of TorchGWAS2 to existing computationally efficient tools such as REGENIE and fastGWA, specifically looking at the wall time and memory usage. For joint analysis of both eyes, we treated the left and right eyes as repeated measurements from a single individual and modelled them as if they were monozygotic twins with the same copy of genetic data, since both eyes share the same genome. We duplicated the genotypes and kinship matrix. The following matrix is an example of duplicated 2 times the kinship matrix for related sample 1 and sample 2 with a kinship coefficient of 0.25:

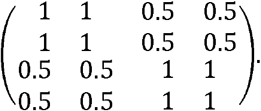

This allows tools such as REGENIE and fastGWA, which are designed for the data with a single measurement per individual, to account for the correlation between two measurements of both eyes. TorchGWAS2 allows repeated measurements for the same person, so we only stacked the phenotypes and covariates files in the long format and kept the kinship matrix genotype files single copied.

### TOPMed metabolomics data

TOPMed Phase 1 circulating metabolomics data included participants from seven cohorts, with metabolite profiling performed at two TOPMed Centralized Omics Resource Core (CORE) laboratories: Baylor-UTHealth Metabolomics Core which used the Metabolon Global Discovery Panel (Metabolon; Morrisville, NC) and Broad-Beth Israel Deaconess Medical Center (Broad-BIDMC). Metabolomics data were harmonized and cataloged as previously described by (Wang, et al., 2025)^12^. The Metabolon platform assayed samples from Genetic Epidemiology of Chronic Obstructive Pulmonary Disease (COPDGene), the Subpopulations and Intermediate Outcomes in Chronic Obstructive Pulmonary Disease Study (SPIROMICS), and Women’s Health Initiative (WHI). The Broad–BIDMC platform assayed samples from the Childhood Asthma Management Program (CAMP), the Genetic Epidemiology of Asthma in Costa Rica study (CRA), the Framingham Heart Study (FHS), the Multi-Ethnic Study of Atherosclerosis (MESA), and WHI. Because a subset of WHI participants had metabolites were measured at both CORE laboratories, metabolites of WHI from different platforms were treated as two independent analytical batches. All included studies were approved by the corresponding institutional review boards, and all participants provided written informed consent.

Metabolites were kept for analysis if they were measured in at least two studies and had a missing rate of less than 50% in each study. For each metabolite, the set of studies in which it was measured was identified, and metabolites were grouped into 11 analysis batches based on the patterns of study availability (Supplementary Table S6). We excluded the study patterns that had fewer than 10 metabolites for the evaluation of tools’ performance on multiple phenotypes. The CAMP study only appeared in small batches, so it was excluded from the primary analysis.

Eventually, 1,023 circulating metabolites measured in 15,565 participants were analyzed with a two-stage procedure. Each metabolite was regressed on fix-effect covariates age, sex, study and the first 11 ancestry PCs. The resulting residuals were rank-normalized and then used as the outcome of following LMM. LMMs on these rank-normalized residuals were adjusted by age, sex, study and the first 11 ancestry PCs as well, following (Wang et al. 2025)^12^. To account for sample relatedness, we incorporated a sparse GRM to model the random effects. The models were consistent with those described in (Wang et al., 2025)^12^. Association analyses were conducted with about 23 million genetic variants with MAF > 0.001 from TOPMed freeze 10 Whole Genome Sequencing data. Detailed variant calling, processing, and quality control were previously described (Taliun et al., 2021)^20^. Further details regarding data processing, and quality control are described on the TOPMed website (https://topmed.nhlbi.nih.gov/topmed-whole-genome-sequencing-methods-freeze-9) and in a common document accompanying each TOPMed study’s dbGaP accession.

### Hardware and software

We ran TorchGWAS2 and benchmarks across different types of platforms. For UK Biobank retinal image data, analyses were performed on a computing server equipped with two AMD EPYC 7352 24-Core processors (48 physical cores, 96 logical threads in total, base clock 2.3 GHz, and 256 MiB L3 cache). GPU-accelerated computations were carried out on one of the eight NVIDIA A100 SXM4 GPUs, each with 80 GB of HBM2 memory, and NVIDIA driver version 580.126.09. For TOPMed Phase I metabolomics data, fastGWA and REGENIE were running on server equipped with dual Intel Xeon Gold 6254 processors, providing a total of 72 logical CPU cores. We also ran GPU-accelerated TorchGWAS2 on Google Cloud with g2-standard-32 with 1 NVIDIA L4 GPU, 32 vCPUs, 128GB random access memory (RAM), and GMMAT on Google Cloud with n1-standard-96 with 96 vCPUs, 360GB RAM.

A benchmark of TorchGWAS2, fastGWA, REGENIE, and GMMAT were conducted, and their performance on runtime, memory usage and costs of cloud computation were evaluated. TorchGWAS2 integrates a stack of scientific computing and genetics libraries across C++, Python and GPU backends. The core C++ module uses MKL or OpenBLAS for BLAS/LAPACK and Eigen 3.4.0 for linear algebra, and SuiteSparse v7.12.1 for sparse matrix operations in the null model fitting step. Genetics data handling relies on PLINK 2.0 with zstd and libdeflate compression. PyTorch serves as the primary computation engine in association testing step. On the CPU, PyTorch is compiled against MKL or OpenBLAS. GPU operations used CUDA libraries cuBLAS bundled with the PyTorch wheel for matrix multiplication. REGENIE v4.1 (https://rgcgithub.github.io/regenie/), fastGWA version 1.94.0beta (https://yanglab.westlake.edu.cn/software/gcta/#fastGWA), and GMMAT v1.4 (https://cran.r-project.org/package=GMMAT) were executed using their publicly available software distributions.

## Supplementary Figures

**Supplementary Figure S1.**
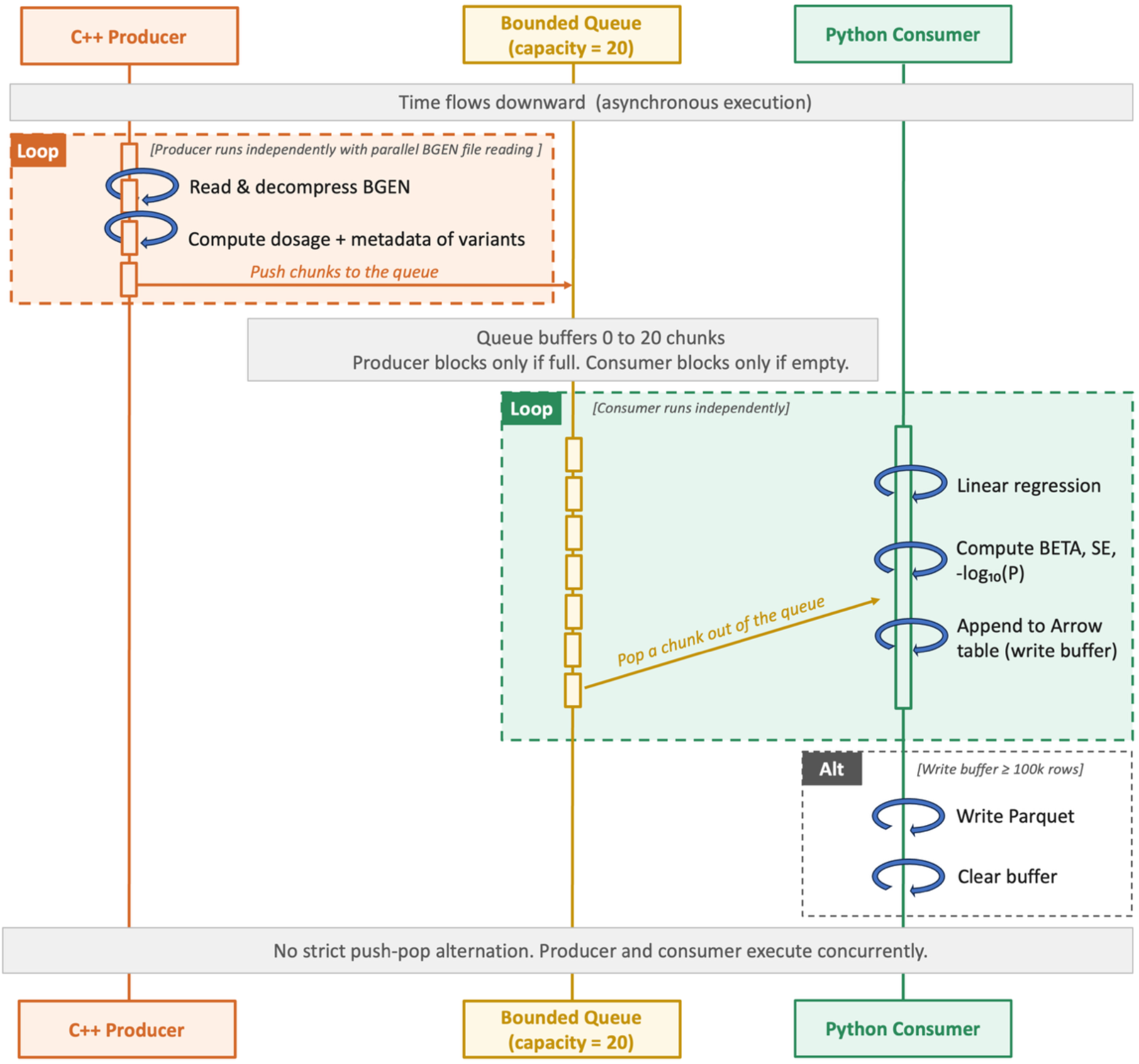
A snapshot of workflows in TorchGWAS2 step 2. Matrix computations in the loop of the Python consumer are accelerated by GPU.

**Supplementary Figure S2.**
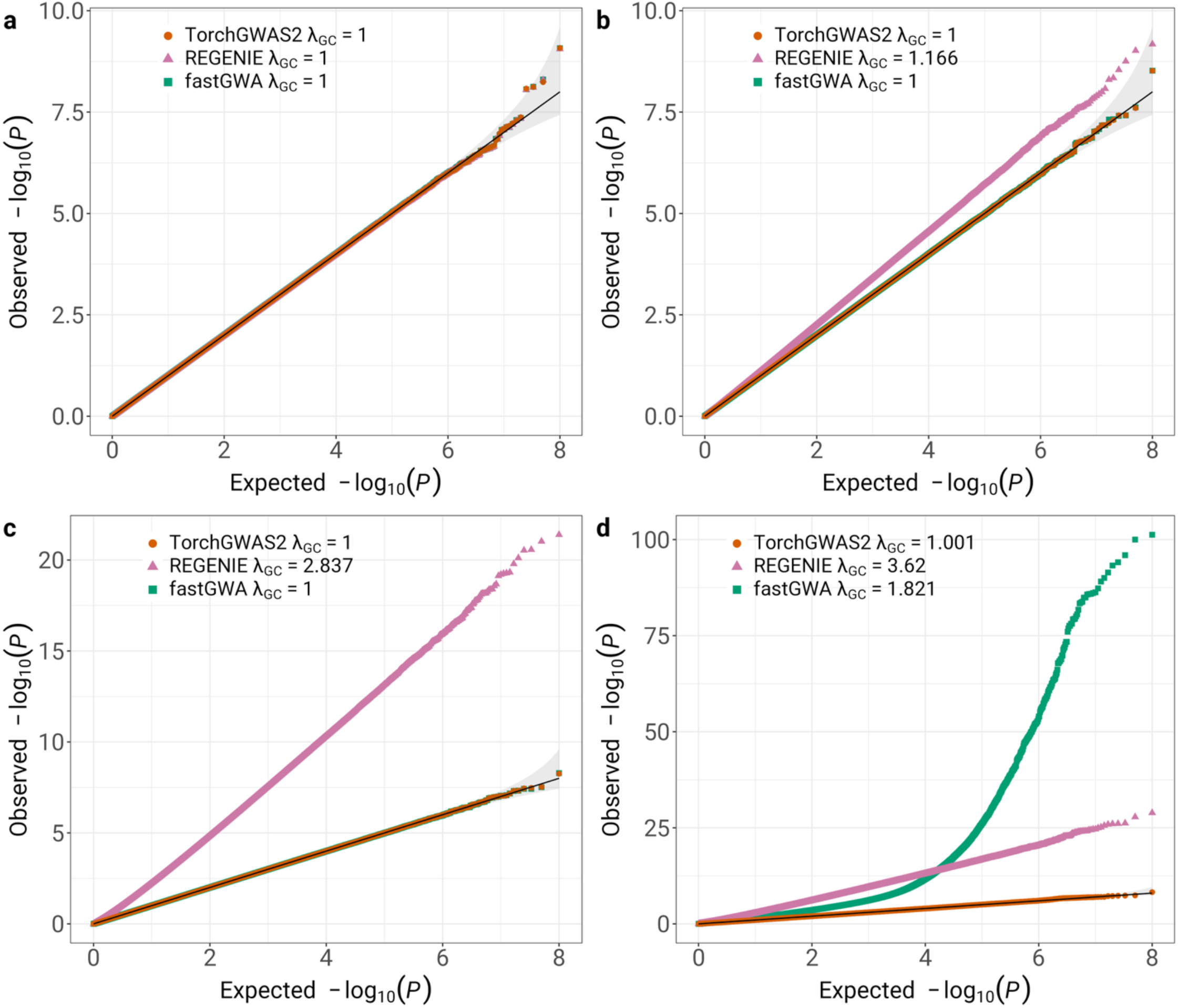
Quantile–quantile (QQ) plots of fastGWA, REGENIE, and TorchGWAS2 *P* values from 100 simulation replicates under the null hypothesis of no genetic effects. **a**, cross-sectional unrelated samples. **b**, cross-sectional related samples. **c**, longitudinal unrelated samples. **d**, longitudinal related samples.

**Supplementary Figure S3.**
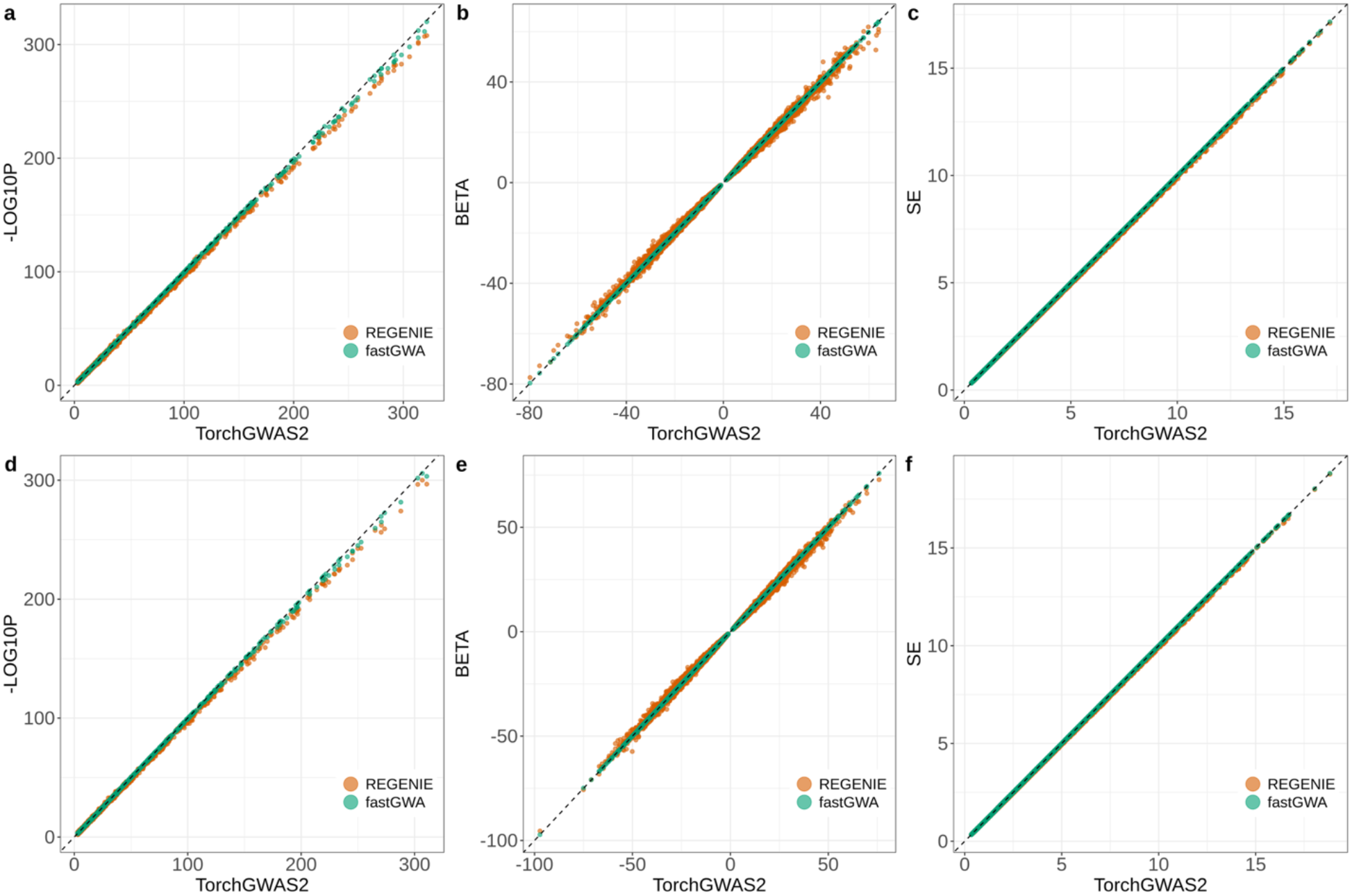
Comparison of *P* values on the −log_10_ scale (**a, d**), effect size estimates (**b, e**), and standard errors (**c, f**) among TorchGWAS2, fastGWA and REGENIE for single-eye analysis combined for all 128 dimensions from UKB retinal image data. **a, b, c**, left eye analysis. **d, e, f**, right eye analysis. The variants in the plots were filtered with MAF > 0.001, and *P <* 0.001. The x-axis represents results from TorchGWAS2, and the y-axis represents results from fastGWA (green) and REGENIE (orange). The dashed black line represents the identity line.

**Supplementary Figure S4.**
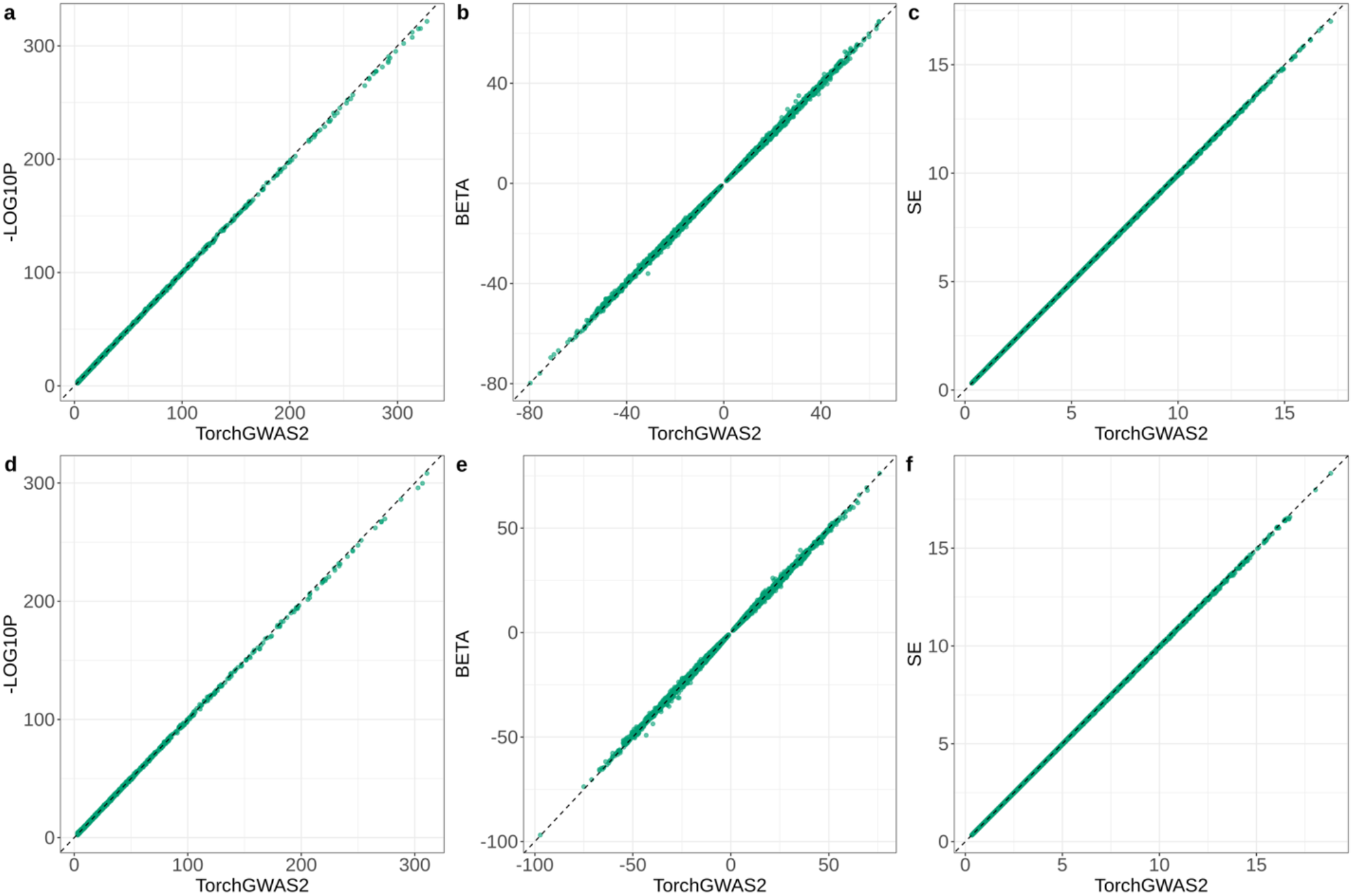
Comparison of *P* values on the −log_10_ scale (**a, d**), effect size estimates (**b, e**), and standard errors (**c, f**) between TorchGWAS2 and BOLT-LMM with the infinitesimal model (BOLT-LMM-inf) for single-eye analysis combined for all 128 dimensions from UKB retinal image data. **a, b, c**, left eye analysis. **d, e, f**, right eye analysis. The variants in the plots were filtered with MAF > 0.001, and *P <* 0.001. The x-axis represents results from TorchGWAS2, and the y-axis represents results from BOLT-LMM. The dashed black line represents the identity line.

**Supplementary Figure S5.**
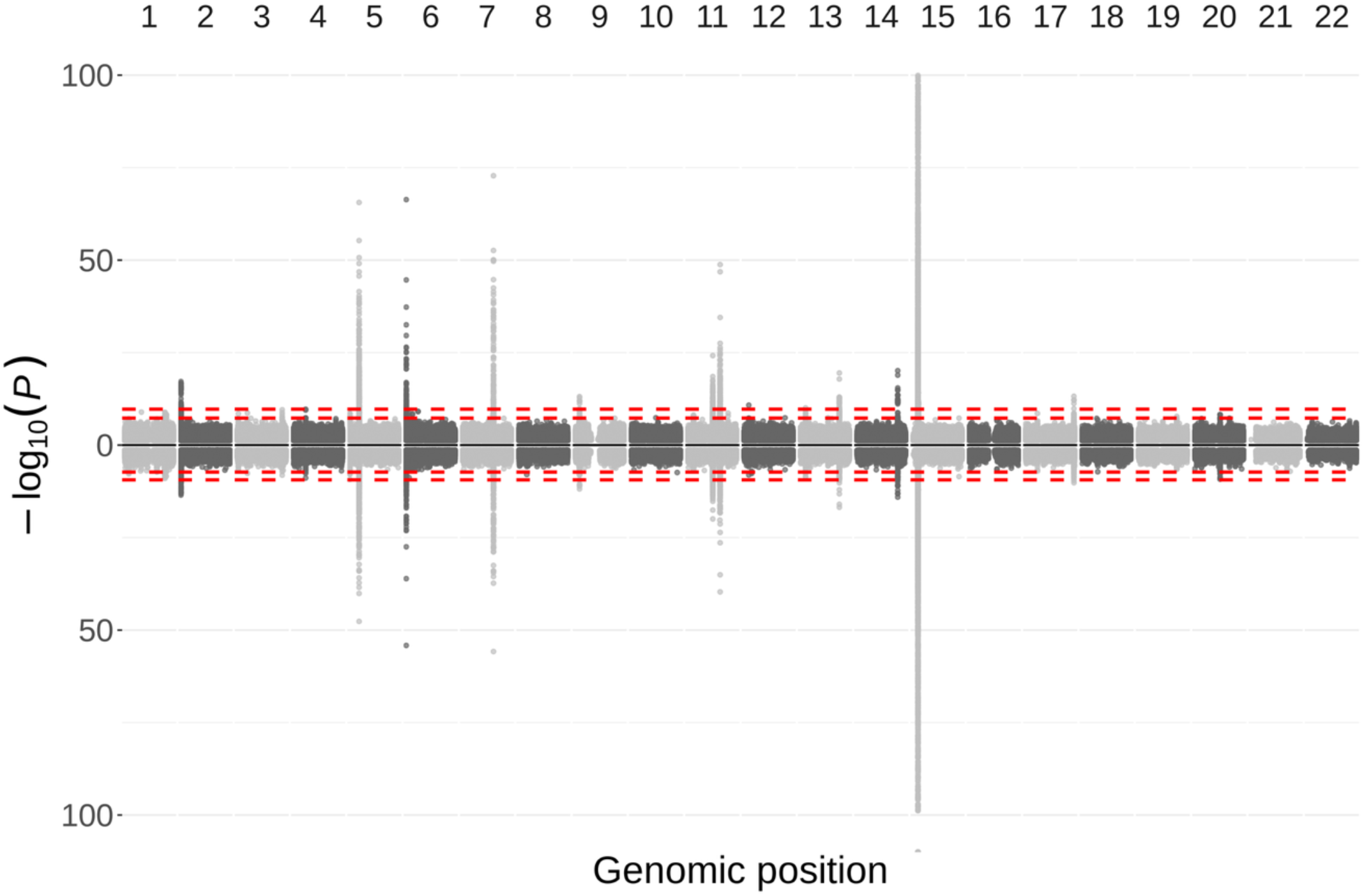
Miami plot of both-eye analysis (top) from TorchGWAS2 and single-eye analysis (bottom) from BOLT-LMM infinitesimal model combined for all 128 dimensions from UKB retinal image data. Minimum *P* value between the left- and right-eye analyses was selected as the single-eye analysis *P* value for each variant. The dashed lines are −log_10_ (5 × 10^−8^) and −log_10_ (5 × 10^−8^/128) for both-eye analyses, and −log_10_ (5 × 10^−8^) and −log_10_ (5 × 10^−8^/256) for single-eye analyses, respectively. Only variants with MAF > 0.001 and *P <* 0.05 are plotted. The plot is cut off at − log_10_(*P*) = 100 for better visualization. The significant regions are provided in the **Supplementary Table S5**.

**Supplementary Figure S6.**
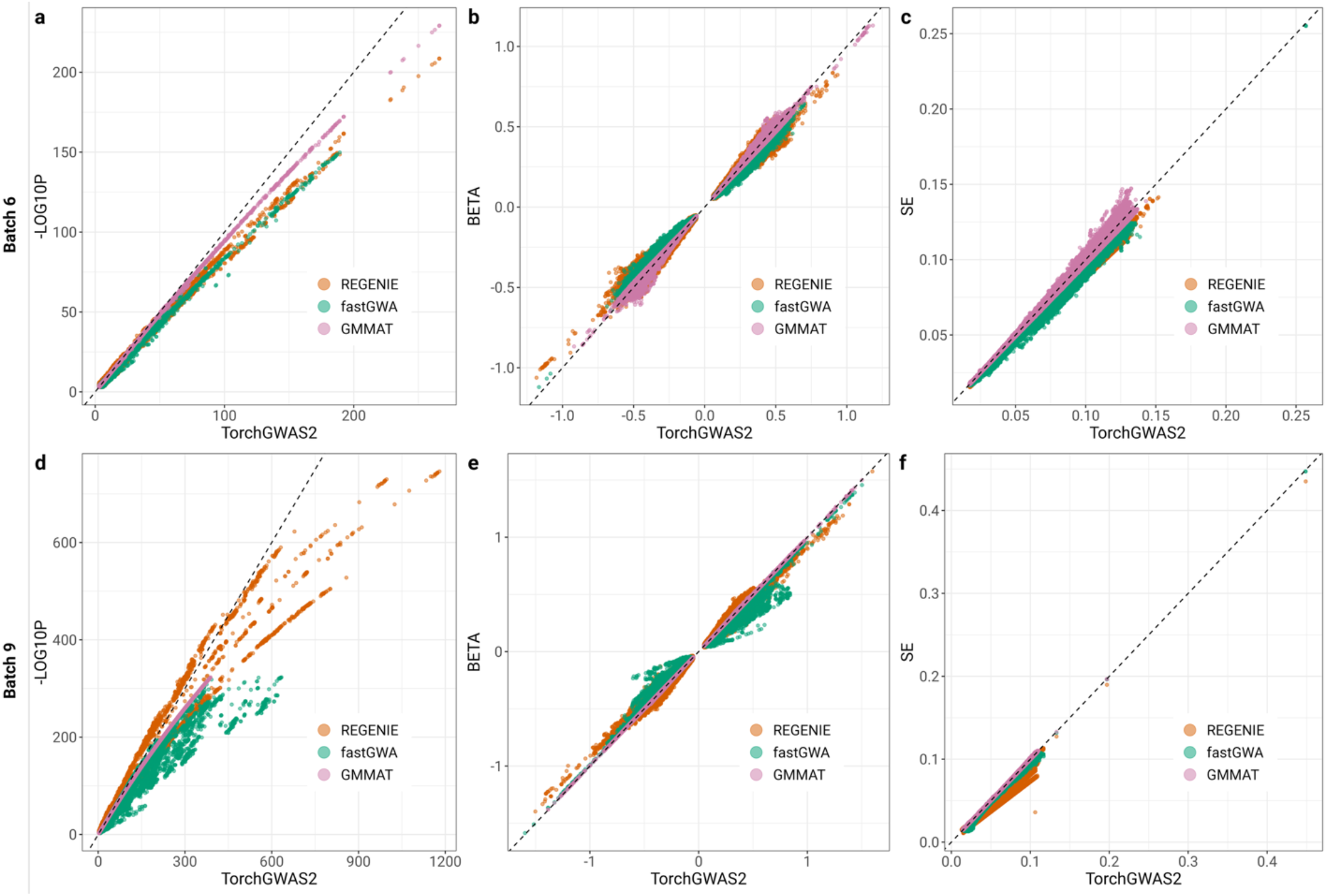
Comparison of *P* values on the −log_10_ scale (**a, d**), effect size estimates (**b, e**), and standard errors (**c, f**) among TorchGWAS2, GMMAT, fastGWA, and REGENIE from TOPMed metQTL analyses batch 6 (**a, b, c**) and batch 9 (**d, e, f**). The variants in the plots were filtered with MAF > 0.005, and *P <* 0.05. The x-axis represents results from TorchGWAS2, and the y-axis represents results from REGENIE (orange), fastGWA (green), and GMMAT (pink). The dashed black line represents the identity line.

**Supplementary Figure S7.**
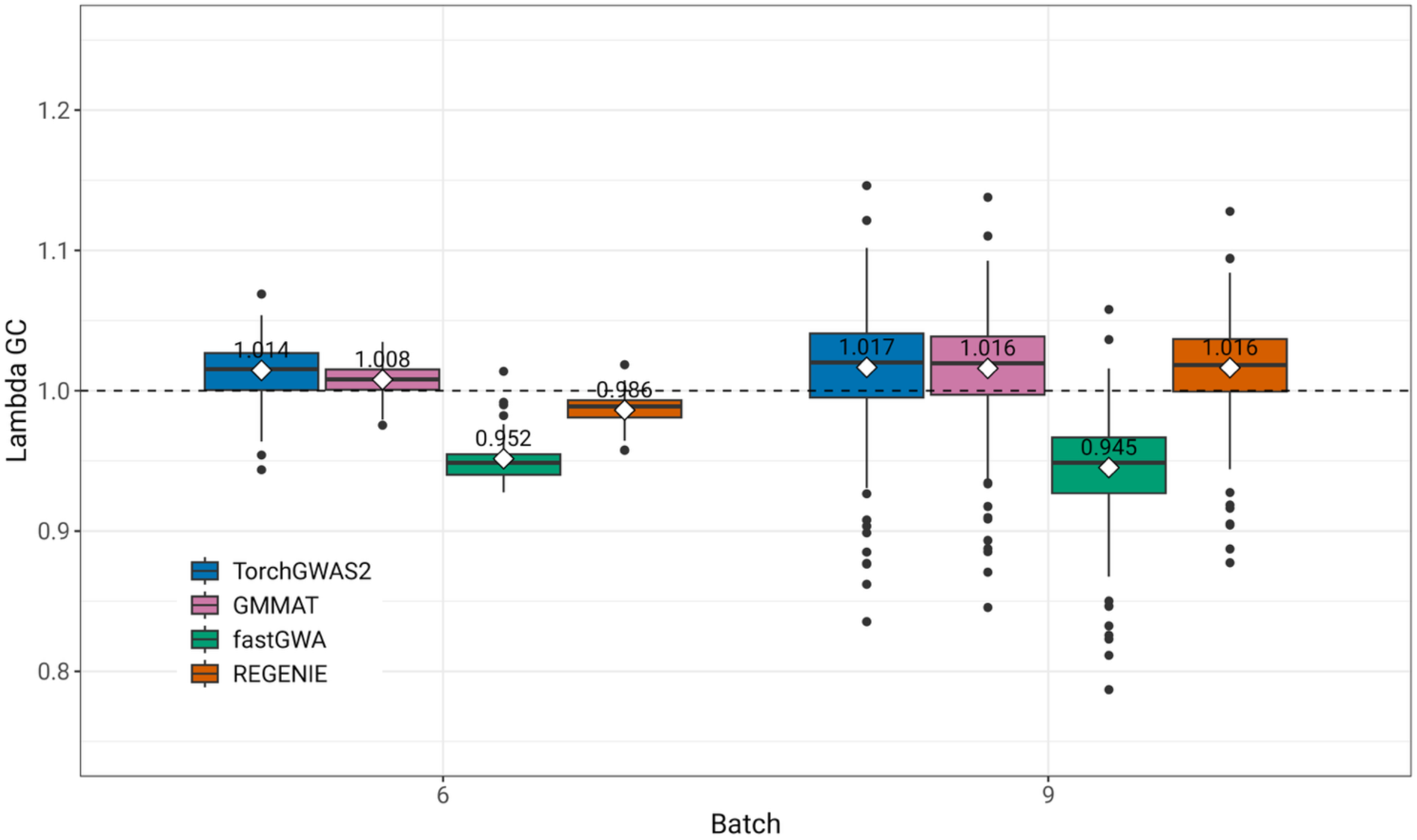
Comparison of genomic inflation factors for TOPMed metQTL analyses from batch 6 and batch 9 among TorchGWAS2, GMMAT, fastGWA, and REGENIE. The white squares show the means of the genomic inflation factors across all metabolites for each batch.

**Supplementary Figure S8.**
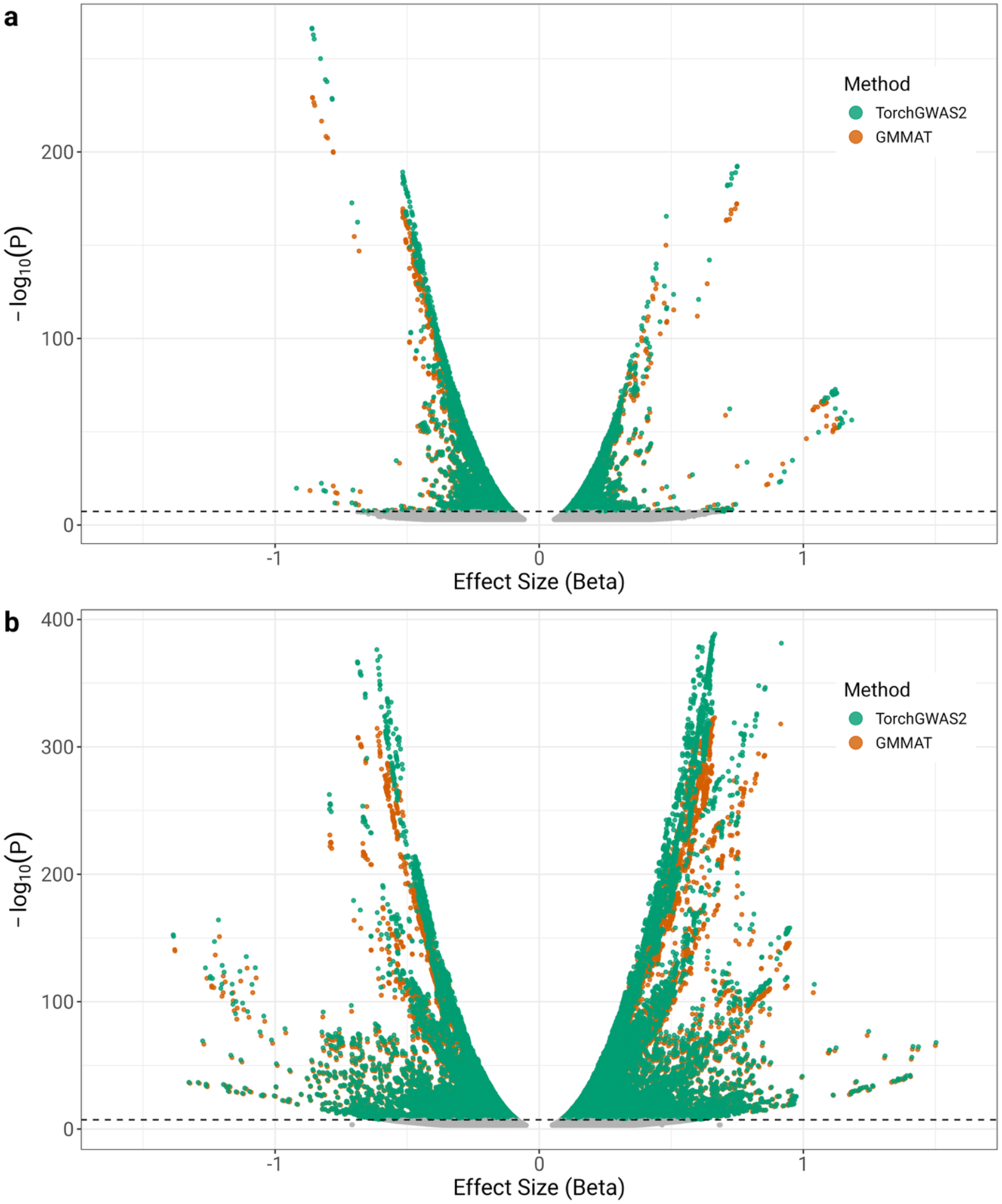
Scatter plot of − log_10_ (*P*) versus effect size of TorchGWAS2 and GMMAT for TOPMed metQTL analyses batch 6 (**a**) and batch 9 (**b**). Variants with MAF > 0.005 and P < 0.05 were plotted. Dashed black line indicates − log_10_(5 × 10^−8^).

## Supplementary Tables

**Supplementary Table S1.** Simulation settings for benchmark TorchGWAS2 against fastGWA and REGENIE.

| Setting | Type I error | Power<br>GWAS of complex traits |  | Power<br>QTL studies |  |
| --- | --- | --- | --- | --- | --- |
|  | Cross-sectional /<br>Longitudinal | Cross-sectional | Longitudinal | Cross-sectional | Longitudinal |
| No. replicates | 1,900<sup>(1)</sup> | 100 | 100 | 100 | 100 |
| **Sample size in complete cases (0% -30% missing rate on phenotypes)** |  |  |  |  |  |
| Unrelated | 20,000 | 20,000 | 20,000 | 400 | 400 |
| Related | 100,000 | 100,000 | 100,000 | 400 | 400 |
| **Genetic architecture** |  |  |  |  |  |
| No. causal variants | 0 | 50 | 50 | 1 | 1 |
| Phenotypic variance explained by causal variants<br>(unrelated / related) | 0% | 2% / 0.5% | 1% / 0.2% | 2% | 1% |

**Supplementary Table S2.**
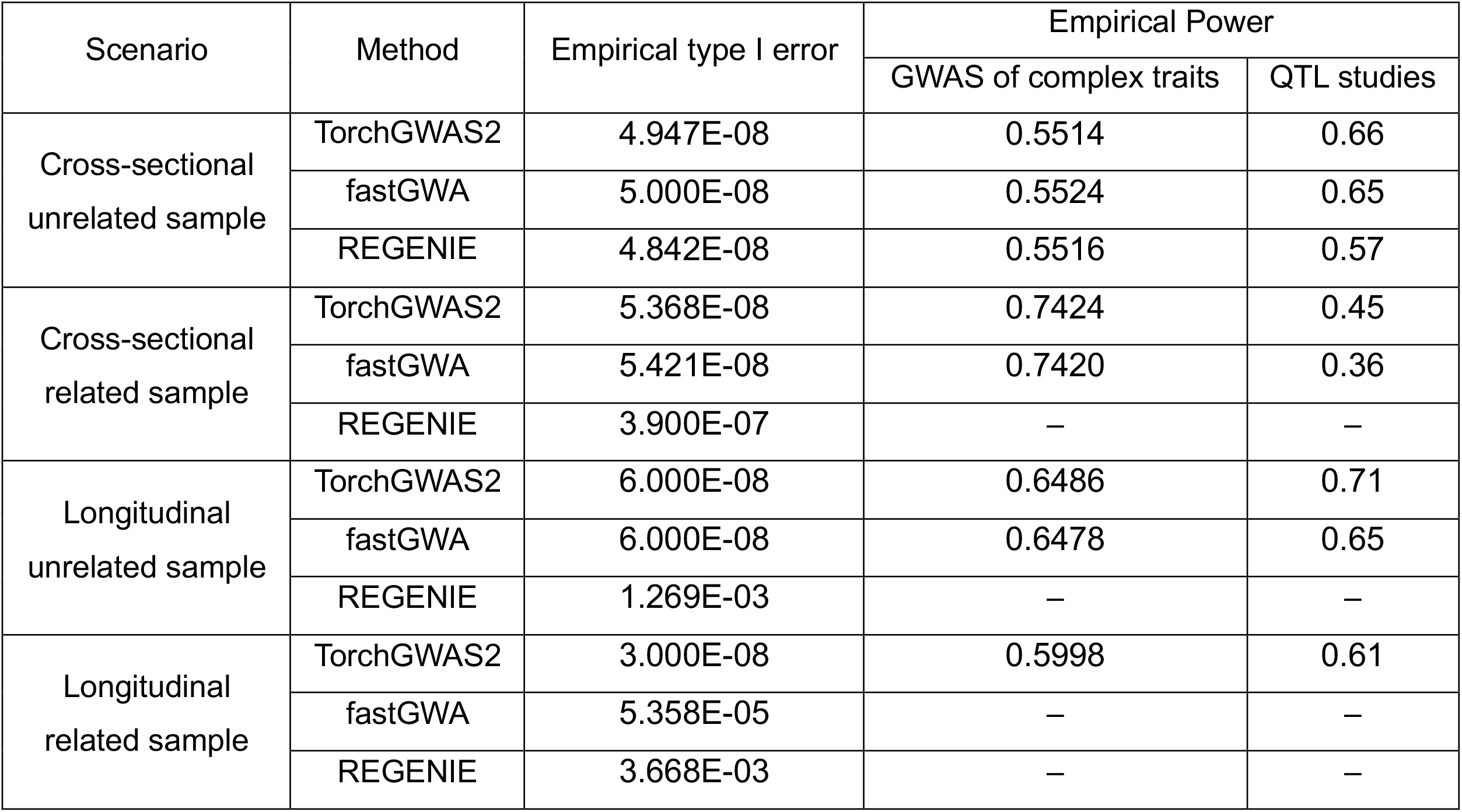
Comparison of empirical type I error rates and power at genome-wide significance level of 5 × 10^−8^ among TorchGWAS2, fastGWA and REGENIE.

**Supplementary Table S3.** Comparison of time and peak memory footprint among TorchGWAS2-GPU, fastGWA and REGENIE for cross-sectional data with 1 million variants and 100,000 related samples.

| Methods | 100 phenotypes | 50 phenotypes | 10 phenotypes | 1 phenotype |
| --- | --- | --- | --- | --- |
| **Runtime** |  |  |  |  |
| TorchGWAS2<br>(1000 variants per batch) | 306 s | 280 s | 182 s | 161 s |
| TorchGWAS2<br>(100 variants per batch) | 364 s | 306 s | 198 s | 176 s |
| fastGWA | 7,977 s | 3,986 s | 811 s | 77 s |
| REGENIE | 5,304 s | 3,128 s | 914 s | 367 s |
| **Peak memory footprint** |  |  |  |  |
| TorchGWAS2<br>(1000 variants per batch) | 26.3 GB | 25.06 GB | 21.9 GB | 22.9 GB |
| TorchGWAS2<br>(100 variants per batch) | 8.0 GB | 6.5 GB | 3.8 GB | 3.5 GB |
| fastGWA | 0.9 GB | 0.9 GB | 0.9 GB | 0.8 GB |
| REGENIE | 3.7 GB | 3.2 GB | 2.8 GB | 2.7 GB |

Notes
- (1) 10,000 variants were used in REGENIE step 1. - (2) All methods used 20 threads for parallel computing.

**Supplementary Table S4.** Benchmark of peak memory footprint in the application of UKB retinal imaging data.

| Benchmark | TorchGWAS2 both eye |  | REGENIE | fastGWA |
| --- | --- | --- | --- | --- |
| Both-eye analysis | CPU runs | GPU runs<br>(CPU / GPU) | CPU Memory Usage | CPU Memory Usage |
| Step 1 | 8.4 GB | – | 3.2 GB |  |
| Step 2 | 12 GB | 11.3 GB / 1.5 GB | 2.0 GB | 1.1 GB |
| Step X | 5.8 GB | – | – |  |
| Left-eye analysis | CPU runs | GPU runs<br>(CPU / GPU) | CPU Memory Usage | CPU Memory Usage |
| Step 1 | 4.5 GB | – | 1.7 GB |  |
| Step 2 | 11.6 GB | 16.0 GB / 1.0 GB | 1.8 GB | 0.46 GB |
| Step X | 5.4 GB | – | – |  |
| Right eye | CPU runs | GPU runs<br>(CPU / GPU) | CPU Memory Usage | CPU Memory Usage |
| Step 1 | 4.5 GB | – | 1.7 GB |  |
| Step 2 | 11.5 GB | 11.2 GB / 1.0 GB | 1.1 GB | 0.46 GB |
| Step X | 6.0 GB | – | – |  |

Notes
(1) all methods used 20 threads for parallel computing. Step X means estimating tuning parameter step in fastGWA and file conversion step in TorchGWAS2.

**Supplementary Table S5.** Significant loci identified in UKB retinal imaging data both-eye analysis and single-eye analysis with TorchGWAS2 and single-eye analysis with BOLT-LMM.

| Region | Lead SNP | CHR | BP | Both-eye<br>*P* values | Single-eye<br>*P* values | Region Start Base Position | Region End Base Position | NearestGene<br>within 500kb window | Both- or<br>single-eye analysis |
| --- | --- | --- | --- | --- | --- | --- | --- | --- | --- |
| 1 | rs11553746 | 2 | 272203 | 6.75E-18 | 3.10E-14 | 30703 | 518055 | *SH3YL1, ACP1* | Shared |
| 2 | rs9838604 | 3 | 181346790 | 2.79E-10 | 7.50E-09 | 181116418 | 181595982 | – | Both-eye only |
| 3 | rs4694845 | 4 | 47368627 | 1.96E-10 | 1.30E-09 | 47292034 | 47617924 | *GABRB1* | Both-eye only |
| 4 | rs16891982 | 5 | 33951693 | 2.65E-66 | 2.20E-48 | 33832433 | 33969628 | *SLC45A2* | Shared |
| 5 | rs12203592 | 6 | 396321 | 4.45E-67 | 7.00E-55 | 204072 | 626889 | *IRF4* | Shared |
| 6 | rs117024920 | 7 | 99446494 | 3.10E-14 | 6.63E-13 | 99250475 | 99691740 | *CPSF4* | Shared |
| 7 | rs80308281 | 7 | 100457578 | 1.56E-73 | 1.60E-56 | 100227188 | 100701451 | *SPACDR* | Shared |
| 8 | rs1408799 | 9 | 12672097 | 7.78E-14 | 1.40E-12 | 12482266 | 12921348 | *LURAP1L-AS1* | Shared |
| 9 | rs72928978 | 11 | 68831364 | 6.44E-25 | 1.10E-20 | 68595250 | 69088752 | *CPT1A, MRPL21* | Shared |
| 10 | rs1847134 | 11 | 89005253 | 1.61E-49 | 2.10E-40 | 88137912 | 89234026 | *CTSC, GRM5* | Shared |
| 11 | rs1684387 | 12 | 13072952 | 1.59E-11 | 8.30E-09 | 13054317 | 13184050 | *FAM234B* | Both-eye only |
| 12 | rs11616792 | 13 | 29165349 | 8.73E-11 | 2.40E-09 | 28941930 | 29366468 | *MTUS2* | Both-eye only |
| 13 | rs9301972 | 13 | 95164550 | 3.20E-20 | 1.60E-17 | 94934930 | 95395373 | *ABCC4* | Shared |
| 14 | rs12896399 | 14 | 92773663 | 7.62E-21 | 8.60E-15 | 92557264 | 92964088 | – | Shared |
| 15 | rs1129038 | 15 | 28356859 | 1.69E-812 | 3.02E-322 | 27786930 | 29429017 | *ENTREP2, APBA2,*<br>*MIR4509-2, HERC2, OCA2* | Shared |
| 16 | rs67050149 | 17 | 79557043 | 6.44E-14 | 6.60E-11 | 79354342 | 79693829 | *RBFOX3* | Shared |

**Notes**
(1) The significance level of both-eye analysis is $5 \times 10^{-8}/128$ , and the significance level of single-eye analysis is $5 \times 10^{-8}/256$ .
(2) Variants were clumped into independent loci if they were in linkage disequilibrium ( $r^2 > 0.001$ ) and located within 250 kb of each.
(3) Single-eye *P* values were generated with BOLT-LMM from Xie *et al.*, 2024<sup>1</sup>.

**Supplementary Table S6.** Summary of metabolomics batches included in the analysis, including the metabolomics platform used, contributing cohort, number of metabolites analyzed, and sample size for each batch in TOPMed metQTL analyses. Metabolomics platforms are Baylor-UTHealth Metabolomics Core which used the Metabolon Global Discovery Panel (Metabolon; Morrisville, NC) and Broad-Beth Israel Deaconess Medical Center (Broad-BIDMC).

| Batch | Metabolomics platform | Contributing cohorts | Metabolites analyzed | Sample size (N) |
| --- | --- | --- | --- | --- |
| 1 | Metabolon | SPIROMICS, WHI | 44 | 2,469 |
| 2 | Broad-BIDMC | WHI, CRA | 73 | 2,778 |
| 3 | Broad-BIDMC | WHI, FHS | 11 | 4,045 |
| 4 | Broad-BIDMC | WHI, FHS, MESA | 14 | 5,043 |
| 5 | Broad-BIDMC | WHI, CRA, FHS | 47 | 5,799 |
| 6 | Broad-BIDMC | WHI, CRA, FHS, MESA | 124 | 6,797 |
| 7 | Metabolon | COPDGene, WHI | 21 | 6,844 |
| 8 | Metabolon | COPDGene, SPIROMICS | 17 | 8,223 |
| 9 | Metabolon | COPDGene, SPIROMICS, WHI | 433 | 8,768 |
| 10 | Metabolon + Broad-BIDMC | COPDGene, WHI, CRA, FHS, SPIROMICS | 93 | 14,567 |
| 11 | Metabolon + Broad-BIDMC | COPDGene, WHI, CRA, FHS, SPIROMICS, MESA | 146 | 15,565 |

## Cohort Description

### COPDGene

The multi-center, NIH-funded COPDGene (also known as the Genetic Epidemiology of COPD Study) includes a study population of more than 10,000 smokers (~1/3 self-reported nonHispanic Black and 2/3 non-Hispanic White participants) and has been characterized with a study protocol including pulmonary function tests, chest CT scans, six-minute walk testing, and multiple questionnaires^2^. DNA was collected for whole genome sequencing at the baseline visit in all participants. A subset of participants had follow-up visits; blood collection for omics, including metabolomics, was performed at the five year follow-up (Phase 2) visit. Five years and ten years after this initial visit a similar protocol was conducted at a follow-up visit. More details are available at http://www.copdgene.org/.

### FHS

The Framingham Heart Study (FHS) is a three-generation, single-site, community-based, ongoing cohort study initiated in 1948 to investigate prospectively the risk factors for CVD^3–5^. It now comprises 3 generations of participants: the original cohort followed since 1948; their Offspring and spouses of the Offspring, followed since 1971; and children from the largest Offspring families enrolled in 2002 (Gen 3). The Original cohort enrolled 5,209 men and women (comprising two-thirds of the adult population in Framingham, MA at that time). Survivors continue to receive biennial examinations. The Offspring cohort comprises 5,124 persons (including 3,514 biological offspring) who have been examined approximately once every 4 years. The Gen 3 cohort contains 4,095 participants.

### MESA

The Multi-Ethnic Study of Atherosclerosis (MESA) is designed to assess subclinical cardiovascular disease and the risk factors that predict cardiovascular disease progression. MESA recruited a diverse, population-based sample of 6,814 asymptomatic men and women aged 45-84 at baseline^6^. MESA included participants who were free of clinical cardiovascular disease at baseline. In terms of self-described race and/or ethnicity, 38 percent of the recruited participants were non-Hispanic White, 28 percent Black or African-American, 22 percent Hispanic, and 12 percent Asian, predominantly of Chinese descent. Participants were recruited from six field centers across the United States: Wake Forest University, Columbia University, Johns Hopkins University, University of Minnesota, Northwestern University and the University of California - Los Angeles.

### WHI

The Women’s Health Initiative (WHI) is a long-term, prospective, multi-center cohort study that investigates post-menopausal women’s health, including strategies to prevent heart disease, breast cancer, colon cancer, and osteoporotic fractures^7^. WHI recruited 161,808 women between 1993 and 1998 at 40 centers across the US (50-79 years at baseline). The study consists of two parts: the WHI Clinical Trial, a randomized clinical trial of hormone therapy, dietary modification, and calcium/Vitamin D supplementation, and the WHI Observational Study, assessing the incidence, risk factors, and interventions related to heart disease, cancer, and osteoporotic fractures. WHI samples sequenced in TOPMed were selected on a case/control basis for stroke and venous thromboembolism.

### CRA

The Genetic Epidemiology of Asthma in Costa Rica (CRA) Cohort recruited 4,245 participants, focusing on children with asthma, their parents, and other pedigree relatives, after screening over 9,180 children enrolled in Costa Rican schools^8^. All probands impacted by asthma completed a protocol including questionnaires, spirometry, methacholine challenge testing (if FEV1 was ≥ 65% of predicted), allergy skin testing, and collection of blood (for plasma, DNA and RNA extraction, and measurement of serum total and allergen-specific IgE) and house dust (for measurement of dust mite/cockroach allergens) samples.

### CAMP

As described previously, the Childhood Asthma Management Program (CAMP) recruited 1,041 children from 5–12 years of age with mild-to-moderate persistent asthma from 1993-95 at eight clinical centers^9^. After initial baseline screening to establish asthma severity, participants were randomized to budesonide dry-powder inhaler (200 μg twice daily), nedocromil metered dose inhaler (8 mg twice daily), or matching placebos for 4–6 years.

### SPIROMICS

The Subpopulations and Intermediate Outcomes in COPD Study (SPIROMICS) is a multi-center observational study of chronic obstructive pulmonary disease (COPD), with 2,982 participants aged 40-80 at baseline in the main study^10^. Most participants had current or former tobacco use for longer than 20 pack-years; control participants without a history of smoking or lung disease were also recruited. Participants participated in deep phenotyping related to COPD, including spirometry and a high-resolution chest CT scan and repeat questionnaires regarding tobacco use, pulmonary symptoms (COPD Assessment Test score), respiratory exacerbations, use of oral corticosteroids and antibiotics, emergency department (ED) visits and hospitalizations. More information is available at https://www.spiromics.org/.

## Study Specific Acknowledgments

### NHLBI TOPMed: Genetic Epidemiology of COPD Study (COPDGene)

This work was supported by NHLBI grants U01 HL089897 and U01 HL089856 and by NIH contract 75N92023D00011. The COPDGene study (NCT00608764) has also been supported by the COPD Foundation through contributions made to an Industry Advisory Committee that has included AstraZeneca, Bayer Pharmaceuticals, Boehringer-Ingelheim, Bristol Myers Squibb, Genentech, GlaxoSmithKline, Johnson & Johnson, Novartis, Pfizer, Regeneron, and Sunovion. A full listing of COPDGene investigators can be found at: http://www.copdgene.org/directory

### NHLBI TOPMed: Framingham Heart Study (FHS)

The Framingham Heart Study (FHS) acknowledges the support of contracts NO1-HC-25195, HHSN268201500001I, 75N92019D00031 and 75N92025D00012 from the National Heart, Lung and Blood Institute, grant supplement R01 HL092577-06S1, and TOPMed X01 HL139389 for this research. We also acknowledge the dedication of the FHS study participants without whom this research would not be possible.

### NHLBI TOPMed: Multi-Ethnic Study of Atherosclerosis (MESA)

Molecular data for the Trans-Omics in Precision Medicine (TOPMed) program was supported by the National Heart, Lung and Blood Institute (NHLBI). Core support including centralized genomic read mapping and genotype calling, along with variant quality metrics and filtering were provided by the TOPMed Informatics Research Center (3R01HL-117626-02S1; contract HHSN268201800002I). Core support including phenotype harmonization, data management, sample-identity QC, and general program coordination were provided by the TOPMed Data Coordinating Center (R01HL-120393; U01HL-120393; contract HHSN268201800001I). We gratefully acknowledge the studies and participants who provided biological samples and data for TOPMed. MESA and the MESA SHARe project are conducted and supported by the National Heart, Lung, and Blood Institute (NHLBI) in collaboration with MESA investigators. Support for MESA is provided by contracts 75N92025D00022, 75N92020D00001, HHSN268201500003I, N01-HC-95159, 75N92025D00026, 75N92020D00005, N01-HC-95160, 75N92020D00002, N01-HC-95161, 75N92025D00024, 75N92020D00003, N01-HC-95162, 75N92025D00027, 75N92020D00006, N01-HC-95163, 75N92025D00025, 75N92020D00004, N01-HC-95164, 75N92025D00028, 75N92020D00007, N01-HC-95165, N01-HC-95166, N01-HC-95167, N01HC-95168, N01-HC-95169, UL1-TR-000040, UL1-TR-001079, UL1-TR-001420, UL1TR001881, DK063491, and R01HL105756. The authors thank the MESA participants and the MESA investigators and staff for their valuable contributions. A full list of participating MESA investigators and institutions can be found at http://www.mesa-nhlbi.org.

### NHLBI TOPMed: SubPopulations and InteRmediate Outcome Measures In COPD Study (SPIROMICS)

The authors thank the SPIROMICS participants and participating physicians, investigators, study coordinators, and staff for making this research possible. More information about the study and how to access SPIROMICS data is available at www.spiromics.org. The authors would like to acknowledge the University of North Carolina at Chapel Hill BioSpecimen Processing Facility (http://bsp.web.unc.edu/) and Alexis Lab (https://www.med.unc.edu/cemalb/facultyresearch/alexislab/) for sample processing, storage, and sample disbursements.

We would like to acknowledge the following current and former investigators of the SPIROMICS sites and reading centers: Neil E Alexis, MD; Wayne H Anderson, PhD; Mehrdad Arjomandi, MD; Igor Barjaktarevic, MD, PhD; R Graham Barr, MD, DrPH; Patricia Basta, PhD; Lori A Bateman, MS; Christina Bellinger, MD; Surya P Bhatt, MD; Eugene R Bleecker, MD; Richard C Boucher, MD; Russell P Bowler, MD, PhD; Russell G Buhr, MD, PhD; Stephanie A Christenson, MD; Alejandro P Comellas, MD; Christopher B Cooper, MD, PhD; David J Couper, PhD; Gerard J Criner, MD; Ronald G Crystal, MD; Jeffrey L Curtis, MD; Claire M Doerschuk, MD; Mark T Dransfield, MD; M Bradley Drummond, MD; Christine M Freeman, PhD; Craig Galban, PhD; Katherine Gershner, DO; MeiLan K Han, MD, MS; Nadia N Hansel, MD, MPH; Annette T Hastie, PhD; Eric A Hoffman, PhD; Yvonne J Huang, MD; Robert J Kaner, MD; Richard E Kanner, MD; Mehmet Kesimer, PhD; Eric C Kleerup, MD; Jerry A Krishnan, MD, PhD; Wassim W Labaki, MD; Lisa M LaVange, PhD; Stephen C Lazarus, MD; Fernando J Martinez, MD, MS; MerryLynn McDonald, PhD; Deborah A Meyers, PhD; Wendy C Moore, MD; John D Newell Jr, MD; Elizabeth C Oelsner, MD, MPH; Jill Ohar, MD; Wanda K O’Neal, PhD; Victor E Ortega, MD, PhD; Robert Paine, III, MD; Laura Paulin, MD, MHS; Stephen P Peters, MD, PhD; Cheryl Pirozzi, MD; Nirupama Putcha, MD, MHS; Sanjeev Raman, MBBS, MD; Stephen I Rennard, MD; Donald P Tashkin, MD; J Michael Wells, MD; Robert A Wise, MD; and Prescott G Woodruff, MD, MPH. The project officers from the Lung Division of the National Heart, Lung, and Blood Institute were Lisa Postow, PhD, and Lisa Viviano, BSN; SPIROMICS was supported by contracts from the NIH/NHLBI (HHSN268200900013C, HHSN268200900014C, HHSN268200900015C, HHSN268200900016C, HHSN268200900017C, HHSN268200900018C, HHSN268200900019C, HHSN268200900020C), grants from the NIH/NHLBI (U01 HL137880, U24 HL141762, R01 HL182622, and R01 HL144718), and supplemented by contributions made through the Foundation for the NIH and the COPD Foundation from Amgen; AstraZeneca/MedImmune; Bayer; Bellerophon Therapeutics; Boehringer-Ingelheim Pharmaceuticals, Inc.; Chiesi Farmaceutici S.p.A.; Forest Research Institute, Inc.; Genentech; GlaxoSmithKline; Grifols Therapeutics, Inc.; Ikaria, Inc.; MGC Diagnostics; Novartis Pharmaceuticals Corporation; Nycomed GmbH; Polarean; ProterixBio; Regeneron Pharmaceuticals, Inc.; Sanofi; Sunovion; Takeda Pharmaceutical Company; and Theravance Biopharma and Mylan/Viatris.

### NHLBI TOPMed: Women’s Health Initiative (WHI)

The WHI program is funded by the National Heart, Lung, and Blood Institute, National Institutes of Health, U.S. Department of Health and Human Services through contracts 75N92021D00001, 75N92021D00002, 75N92021D00003, 75N92021D00004, 75N92021D00005. We acknowledge following WHI investigators.

### Program Office

(National Heart, Lung, and Blood Institute, Bethesda, Maryland) Jared Reis and Candice Price. **Clinical Coordinating Center**: (Fred Hutchinson Cancer Center, Seattle, WA) Garnet Anderson, Charles Kooperberg, and Holly Harris. **Steering Committee**: (Fred Hutchinson Cancer Center) Marian Neuhouser – Committee Chair; (Fred Hutchinson Cancer Center) Garnet Anderson; (University of California, Davis) Lorena Garcia; (Wake Forest University) Lindsay Reynolds; (University at Buffalo) Amy Millen; (University at Buffalo) Jean Wactawski-Wende; (Fred Hutchinson Cancer Center) Holly Harris; (University of Massachusetts) Brian Silver; (University of Tennessee Health Center) Karen Johnson; (Stanford Prevention Research Center) Marcia L. Stefanick; (The Ohio State University) Electra Paskett; (Wake Forest University School of Medicine) Mara Vitolins.

